# Frequency and machine learning predictors of depressive, anxiety, obsessive-compulsive symptoms, and suicidal ideation among university students

**DOI:** 10.1101/2023.01.29.23285151

**Authors:** Nicola Meda, Susanna Pardini, Paolo Rigobello, Francesco Visioli, Caterina Novara

## Abstract

**Introduction:** Prospective studies on mental health of students showed that young adults enroled in university are affected by poorer mental health than other working peers or adults, and this condition is responsible for a large proportion of disability-adjusted life-years

**Methods:** We enrolled 1388 students at the baseline (and 557 completed follow-up after six months) who reported their demographic information and completed self-report questionnaires on depressive, anxiety and obsessive-compulsive symptoms. We applied multiple regression modelling and supervised machine learning to evince associations and predict the risk factors of poorer mental health at baseline and follow-up

**Results:** Approximately one out of five students reported severe depressive symptoms and/or suicidal ideation. An association of economic worry with depression was evidenced at the beginning of the study (when there was a high frequency of worry OR = 3.11 [1.88 – 5.15]) and during follow-up. Supervised machine learning exhibited high accuracy in predicting the students who maintained well-being (balanced accuracy = 0.85) or absence of suicidal ideation, but its performance was almost null in identifying those whose symptoms worsened.

**Conclusions:** Students’ severe mental health problems are reaching worrying percentages, and few demographic factors can be leveraged to predict poor mental health outcomes. Further research including people with lived experience is crucial to assess students’ needs and improve the prediction of those at risk of developing worse symptoms.

## Introduction

Students’ mental health has attracted much more research after COVID-19-related lockdowns than ever before (Arsandaux et al., 2021; Bennett et al., 2022; Gestsdottir et al., 2021; Hernández-Torrano et al., 2020; Nuñez et al., 2022; Robinson et al., 2022; Villani et al., 2021; Voltmer et al., 2021; Yamamoto et al., 2022). Prospective studies reported that young adults enrolled in university were affected by symptoms of poorer mental health than other working peers or adults well before restrictive measures were implemented (Blanco et al., 2008; Granieri et al., 2021); In fact, as previously reported, higher education is a risk factor for depressive symptoms, anxiety symptoms, and suicidal thoughts (Karyotaki et al., 2020). In addition, early adulthood is also a crucial transition phase, accompanied by the pursuit of self-efficacy and economic independence, higher education, and new social needs. The Global Burden of Disease Study also evidenced that disability-adjusted life-years are at their highest in young adulthood (“Global, regional, and national burden of 12 mental disorders in 204 countries and territories, 1990–2019,” 2022), in accordance with epidemiological findings that indicate this period of life as the typical age of onset of most mental disorders (Solmi et al., 2021). Until now, research on students’ mental health pinpointed that university support services, where in place, probably had little impact on the general well-being of the student population (Barnett et al., 2021; Eisenberg et al., 2011) for a variety of reasons. Namely, low rates of the number of counselors per student (generating long waitlists – (Blanco et al., 2008; Cohen et al., 2022; Lueck and Poe, 2021)) high levels of personal stigma hindering help-seeking behaviors (Eisenberg et al., 2009), and few systemic interventions to address the determinants of (poor) mental health and prompt an early intervention to assess and treat the symptoms before they develop into a full-blown mental disorder (Upsher et al., 2022) or treat effectively the students with chronic symptoms of mental disorders (Zivin et al., 2009).

Although essential to tailor population specific psychological interventions (Barnett et al., 2021), little prospective evidence has been published regarding the prediction of mental health problems in university students (Ebert et al., 2019; Suldo et al., 2011; Tyssen et al., 2008). A thorough assessment of the needs of the student population and their psychological symptoms (Kitzrow, 2009) – leading to faster and more reliable diagnosis workups - is the first step in a pragmatic approach to design and evaluate the efficacy of specific mental health interventions. However, some studies investigated students’ mental health through variables strictly related to generalized anxiety disorder and depressive disorders, with little indulging in assessing demographic or other determinants of psychological well-being (Sheldon et al., 2021).

To provide better services, prospective studies would be helpful to design that evaluate risk factors at the start of the study (ideally at university/college enrolment) that help predict those students expected to need help in the short, medium, and long term. Several contributions regarding the role of the COVID-19 pandemic evidenced (Meda et al., 2021; Nomura et al., 2022; Weber et al., 2022) that such studies can be (i) conducted with little or no specific funding and (ii) they can help better understand the mental disorder development trajectory in a population at high risk of distress and maladjustment.

This work is part of a larger prospective study to evaluate the depressive, anxiety, eating, and obsessive-compulsive symptoms of students and their determinants. Here, we report prospective data collected at enrollment and after six months, of hundreds of Italian university students who completed several questionnaires investigating mental health problems. The aims of this study were to establish the percentage of students suffering from severe symptoms common to depression, anxiety, obsessive-compulsive disorder, and suicidal ideation at baseline; to elucidate the socio-demographic variables (e.g., gender, financial situation) associated with poor mental health at baseline; and to evaluate the risk factors (e.g., depressive symptoms at enrollment) that dent the chances of symptoms improvement or otherwise make symptom aggravation probable. In addition, we wanted to establish which parsimonious self-report measures could explain a substantial part of current and future mental health problems. To do so, we employed binomial regression modelling to evince the association between demographic-individual factors and severe depressive, anxiety, obsessive-compulsive symptoms, and suicidal ideation. Lastly, we adopted a supervised machine learning approach to determine which factors at baseline could be used to predict a change in symptoms (new onset, stability, or improvement) after six months.

## Materials and Methods

### Study cohort

All procedures described in this research were approved by the University of Padova Psychology Ethical Committee (Area 17 - ECOS: Eating, Compulsive, and Obsessive Symptoms in Young Adults Protocol Ref. 3005) under the latest version of the Declaration of Helsinki. A significant part of the procedures described herein was also employed in two previous articles derived from the same protocol (Meda et al., 2021; Novara et al., 2022). Participants provided written informed consent to the study. Recruitment took place in Padova, Italy, between October 2019 and October 2020. From October 2019 to March 2020, at the start of different teaching classes, we presented the aims of the prospective study in person and the attending students were handed out a card containing an URL. By accessing the URL, students could provide their informed consent and participate in the study.

From March 2020 onwards, due to COVID-related restrictions, the study was described via pre-recorded videos or remote presentations at the beginning of teaching classes. Throughout the slide show, students had the possibility to copy the URL redirecting to the informed consent. Enrollment and follow-up continued throughout 2022. After the acquisition of informed consent, participants were required to complete a demographic schedule, as well as self-reported questionnaires on mental health on the REDCap (Harris et al., 2009) web application. Every six months since enrollment (for a total of six cross-sectional evaluations), the participants were automatically contacted by email and asked to participate in another data collection. A total of 1,902 students agreed to participate in the study (21.3% response rate, approximately 9000 students were invited to participate) at the first cross-sectional. Of this pool, 1388 participants matched the target population characteristics (Italian native-speaker students aged 18–30); participants were excluded if non-native speakers outside the previous age range or partially completed the questionnaires. No other inclusion or exclusion criteria were applied. Students who participated in the study are (or were) enroled in Medicine and Surgery, Psychology/Mental Health & Neuroscience, Pharmacy and other health sciences, STEM sciences, Arts & Humanities, Law, Economics, and Political Sciences. At the second cross-sectional (six months after the first questionnaire administration), 768 students agreed to participate in the study for the second time. Of these students, 557 were deemed eligible according to the previously outlined criteria. Sample characteristics at first cross-sectional and comparison between the subsample of dropouts to second-time participants are reported in Tables 1 and 2, respectively.

**Table 1.**
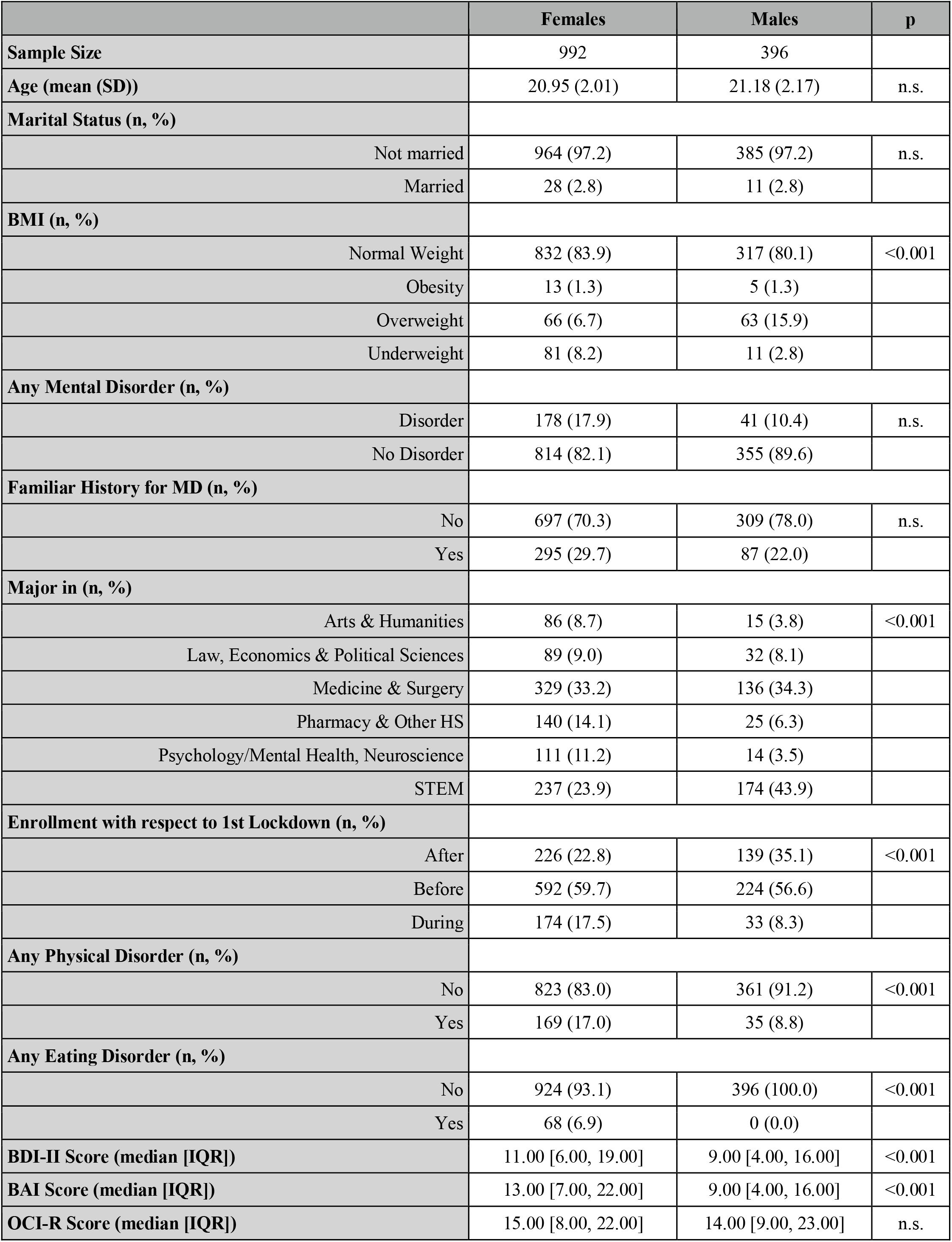
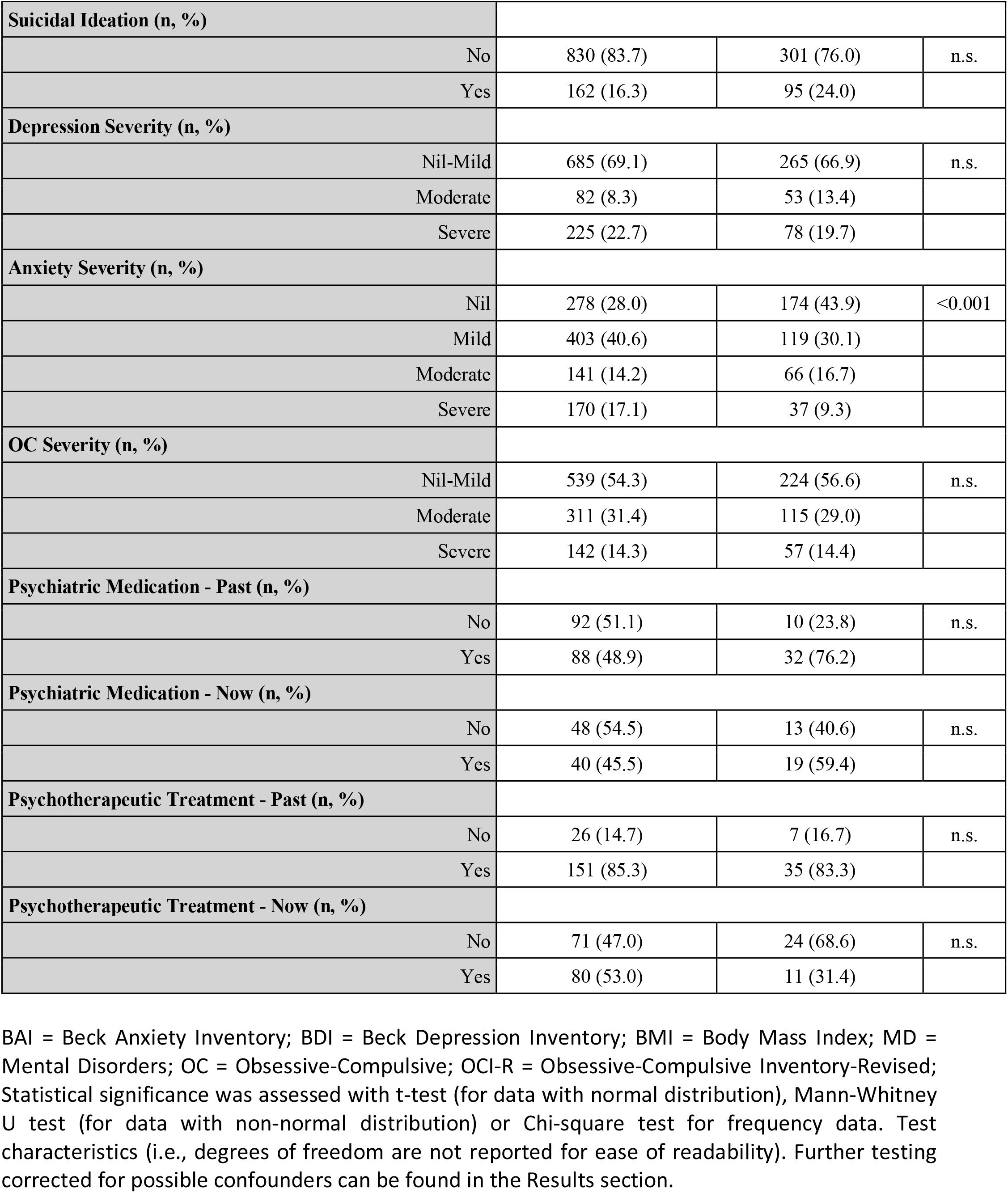
Sample characteristics at baseline.

|  | Females | Males | p |
| --- | --- | --- | --- |
| **Sample Size** | 992 | 396 |  |
| **Age (mean (SD))** | 20.95 (2.01) | 21.18 (2.17) | n.s. |
| **Marital Status (n, %)** |  |  |  |
| Not married | 964 (97.2) | 385 (97.2) | n.s. |
| Married | 28 (2.8) | 11 (2.8) |  |
| **BMI (n, %)** |  |  |  |
| Normal Weight | 832 (83.9) | 317 (80.1) | <0.001 |
| Obesity | 13 (1.3) | 5 (1.3) |  |
| Overweight | 66 (6.7) | 63 (15.9) |  |
| Underweight | 81 (8.2) | 11 (2.8) |  |
| **Any Mental Disorder (n, %)** |  |  |  |
| Disorder | 178 (17.9) | 41 (10.4) | n.s. |
| No Disorder | 814 (82.1) | 355 (89.6) |  |
| **Familiar History for MD (n, %)** |  |  |  |
| No | 697 (70.3) | 309 (78.0) | n.s. |
| Yes | 295 (29.7) | 87 (22.0) |  |
| **Major in (n, %)** |  |  |  |
| Arts & Humanities | 86 (8.7) | 15 (3.8) | <0.001 |
| Law, Economics & Political Sciences | 89 (9.0) | 32 (8.1) |  |
| Medicine & Surgery | 329 (33.2) | 136 (34.3) |  |
| Pharmacy & Other HS | 140 (14.1) | 25 (6.3) |  |
| Psychology/Mental Health, Neuroscience | 111 (11.2) | 14 (3.5) |  |
| STEM | 237 (23.9) | 174 (43.9) |  |
| **Enrollment with respect to 1st Lockdown (n, %)** |  |  |  |
| After | 226 (22.8) | 139 (35.1) | <0.001 |
| Before | 592 (59.7) | 224 (56.6) |  |
| During | 174 (17.5) | 33 (8.3) |  |
| **Any Physical Disorder (n, %)** |  |  |  |
| No | 823 (83.0) | 361 (91.2) | <0.001 |
| Yes | 169 (17.0) | 35 (8.8) |  |
| **Any Eating Disorder (n, %)** |  |  |  |
| No | 924 (93.1) | 396 (100.0) | <0.001 |
| Yes | 68 (6.9) | 0 (0.0) |  |
| **BDI-II Score (median [IQR])** | 11.00 [6.00, 19.00] | 9.00 [4.00, 16.00] | <0.001 |
| **BAI Score (median [IQR])** | 13.00 [7.00, 22.00] | 9.00 [4.00, 16.00] | <0.001 |
| **OCI-R Score (median [IQR])** | 15.00 [8.00, 22.00] | 14.00 [9.00, 23.00] | n.s. |

|  |  |  |  |  |
| --- | --- | --- | --- | --- |
| **Suicidal Ideation (n, %)** |  |  |  |  |
|  | No | 830 (83.7) | 301 (76.0) | n.s. |
|  | Yes | 162 (16.3) | 95 (24.0) |  |
| **Depression Severity (n, %)** |  |  |  |  |
|  | Nil-Mild | 685 (69.1) | 265 (66.9) | n.s. |
|  | Moderate | 82 (8.3) | 53 (13.4) |  |
|  | Severe | 225 (22.7) | 78 (19.7) |  |
| **Anxiety Severity (n, %)** |  |  |  |  |
|  | Nil | 278 (28.0) | 174 (43.9) | <0.001 |
|  | Mild | 403 (40.6) | 119 (30.1) |  |
|  | Moderate | 141 (14.2) | 66 (16.7) |  |
|  | Severe | 170 (17.1) | 37 (9.3) |  |
| **OC Severity (n, %)** |  |  |  |  |
|  | Nil-Mild | 539 (54.3) | 224 (56.6) | n.s. |
|  | Moderate | 311 (31.4) | 115 (29.0) |  |
|  | Severe | 142 (14.3) | 57 (14.4) |  |
| **Psychiatric Medication - Past (n, %)** |  |  |  |  |
|  | No | 92 (51.1) | 10 (23.8) | n.s. |
|  | Yes | 88 (48.9) | 32 (76.2) |  |
| **Psychiatric Medication - Now (n, %)** |  |  |  |  |
|  | No | 48 (54.5) | 13 (40.6) | n.s. |
|  | Yes | 40 (45.5) | 19 (59.4) |  |
| **Psychotherapeutic Treatment - Past (n, %)** |  |  |  |  |
|  | No | 26 (14.7) | 7 (16.7) | n.s. |
|  | Yes | 151 (85.3) | 35 (83.3) |  |
| **Psychotherapeutic Treatment - Now (n, %)** |  |  |  |  |
|  | No | 71 (47.0) | 24 (68.6) | n.s. |
|  | Yes | 80 (53.0) | 11 (31.4) |  |

BAI = Beck Anxiety Inventory; BDI = Beck Depression Inventory; BMI = Body Mass Index; MD = Mental Disorders; OC = Obsessive-Compulsive; OCI-R = Obsessive-Compulsive Inventory-Revised; Statistical significance was assessed with t-test (for data with normal distribution), Mann-Whitney U test (for data with non-normal distribution) or Chi-square test for frequency data. Test characteristics (i.e., degrees of freedom are not reported for ease of readability). Further testing corrected for possible confounders can be found in the Results section.

**Table 2.**
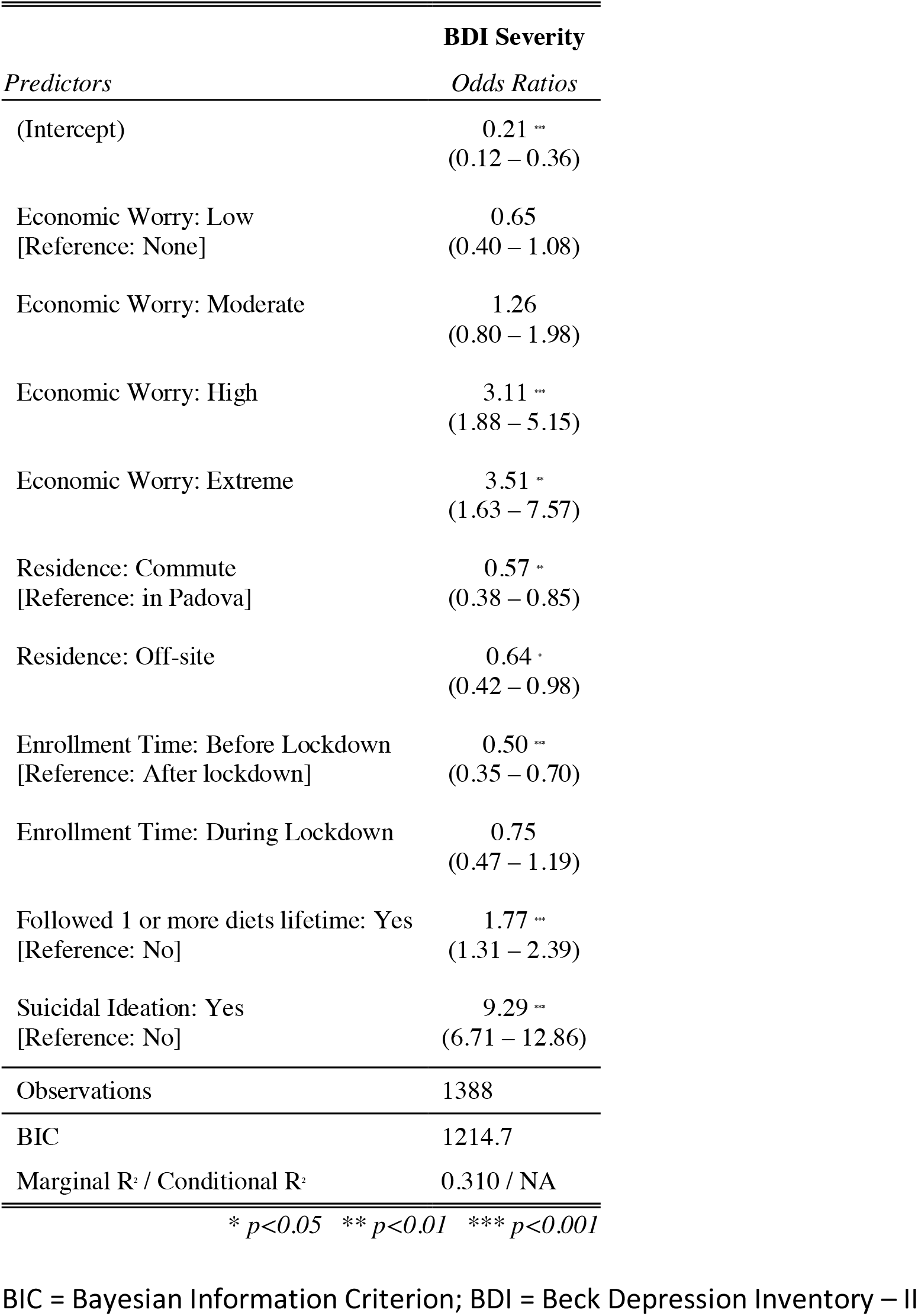
Regression model for severe depressive symptoms.

### Measures

Participants were asked to report their demographic information, years of education, enrollment status, if they had ever been diagnosed with a mental health condition or other medical disorder, illness, or disease, or if they had a family history of mental health problems. Further questions investigated lifestyle habits, such as diet regimen, prescription of medications, past / current psychotherapy, and frequency of worry about their economic/financial conditions. The latter information was assessed with a 5-point Likert scale, where 0 = never worried (nil); 1 = rarely worried (mild); 2 = sometimes worried (moderate); 3 = frequently worried (high); 4 = always worried (extreme). To specifically design this measure, we defined the item similarly to the Worry Domains Questionnaire (Tallis et al., 1992).

Five self-report questionnaires were administered: Beck Depression Inventory II ((Beck et al., 2011; Sica and Ghisi, 2007) BDI-II) to assess depression characteristics (in our sample, this measure maintained a high internal consistency, Cronbach’s α = 0.91 [range: .91 – .92]). The Beck Anxiety Inventory ((Sica and Ghisi, 2007; Steer and Beck, 1997) BAI) for measuring the physical and cognitive symptoms of anxiety (Cronbach’s α=0.92 [range: .91 – .93]). The Obsessive–Compulsive Inventory-Revised ((Abramowitz and Deacon, 2006; Sica et al., 2009) OCI-R), the Eating Disorder Inventory – 3 ((Garner, 2004; Giannini et al., 2008) EDI-3) and the Eating Habits Questionnaire ((Gleaves et al., 2013; Novara et al., 2017) EHQ) [52–71]. For ease of readability and consistency, in this article we will discuss only the results related to depressive, anxious, and obsessive symptoms, as well as suicidal ideation.

### Statistical analysis

Anonymised data were downloaded from the REDCap platform and curated using RStudio – R version 4.1.2 (*R Core Team (2019)*). In this work, we investigated the variables associated with depressive, anxious, obsessive-compulsive symptoms and suicidal ideation at baseline, as well as baseline predictors of mental health outcomes at the first follow-up (six months after baseline). We analyse factors associated with more severe symptoms of depression, anxiety, and obsessive-compulsive behaviour. For the purpose of binomial regression analysis and using random forest algorithms, we labelled the participants with severe symptomatology (=1) or without it (=0), as measured with the self-report measures. The reasons for employing a stringent cut-off are two-fold: first, putative cut-offs for distinguishing nil, mild, and moderate severity of symptoms are less reliable; second, the number of possible changes (and stability) in the severity is four times the number of possible changes with a single cut-off (see also below in the *Prospective Assessment* section). The cutoffs applied were based on the validation procedure of the single psychometric tools. They are as follows: 95^th^ percentile of the BDI-II score (i.e., female score above 20; male score above 19); Score above 26 for the BAI scale (derived from the validation study, for both genders); Score above 28 for the OCI-R scale (derived from the validation study based on the AUC, for both genders). Suicidal ideation was measured with item 9 of the BDI scale (Desseilles et al., 2012; Green et al., 2015).

First, we implemented a regression modeling approach to evaluate the weight of several independent variables on the outcomes of interest, while covarying for possible confounders. In this case, we used a binomial distribution to model the data and show the association between severe symptoms and independent variables. Here we report only the models with the lowest Bayesian Information Criterion (BIC —an index of model fitting: the lower the BIC, the lower the variance left unexplained by the model, the better the fit), which were identified through a stepwise selection approach (Raftery, 1995). We report the β estimate for each variable of the model with the lowest BIC. The estimation of a variable represents the importance of that variable in changing the questionnaire scores.

### Prospective assessment

Then, to identify the predictors of severity changes from baseline to six months after enrollment, we implemented random forest algorithms (supervised machine learning), which extracted from the dataset the independent variables at baseline that played a statistically significant role in determining the change in the symptomatology (i.e., stable “well-being” – not severe at baseline and after six months; stable “severity” – severe both at baseline and after six months; improvement – from severe to not severe symptoms in six months; worsening – from not severe to severe in six months). The algorithms were first trained using 80% of the dataset, and then tested to assess their performance on the remaining 20% of the dataset. Lastly, we evaluated the weight (β estimate) of the independent variables extracted by the algorithm by applying binomial regression modelling. The random forest explainer package (Paluszynska et al., 2020) aided the visualisation of random forest characteristics and predictive performance.

## Results

A total of 1388 students participated in the study (992 – 71.4% females). Some relevant statistically significant differences between the genders emerged with an uncorrected direct comparison approach (Table 1). Females were more likely to be underweight (8.2%) than males (2.8%), who, on the other hand, reported being overweight more frequently (15.19%) than females (6.7%). Most of the participants were recruited before the first COVID-19-related lockdown in Italy (that lasted from the second week of March 2020 to the first week of May 2020).

Females were also more likely to suffer from any physical disorder (17%) or eating disorder (6.9%) than males (8.8% and 0% respectively). Moreover, median scores for depressive and anxiety symptomatology were higher in females than males. However, the frequency of severe symptoms of depression was not different between the sexes: only the frequency of severe symptoms of anxiety was higher in women than in men. Noteworthily, the frequencies of suicidal ideation were not different between the genders.

Importantly, there were no statistically significant differences in the frequency of self-reported, clinician-diagnosed mental health condition diagnosis between genders or familiarity with mental disorders.

For quick comparison (uncorrected for confounder variables), we report in Figure 1 the percentage of severe symptoms (of depression, anxiety, or obsessive-compulsive symptomatology), as well as the frequency of suicidal ideation by gender and field of study.

**Figure 1.**
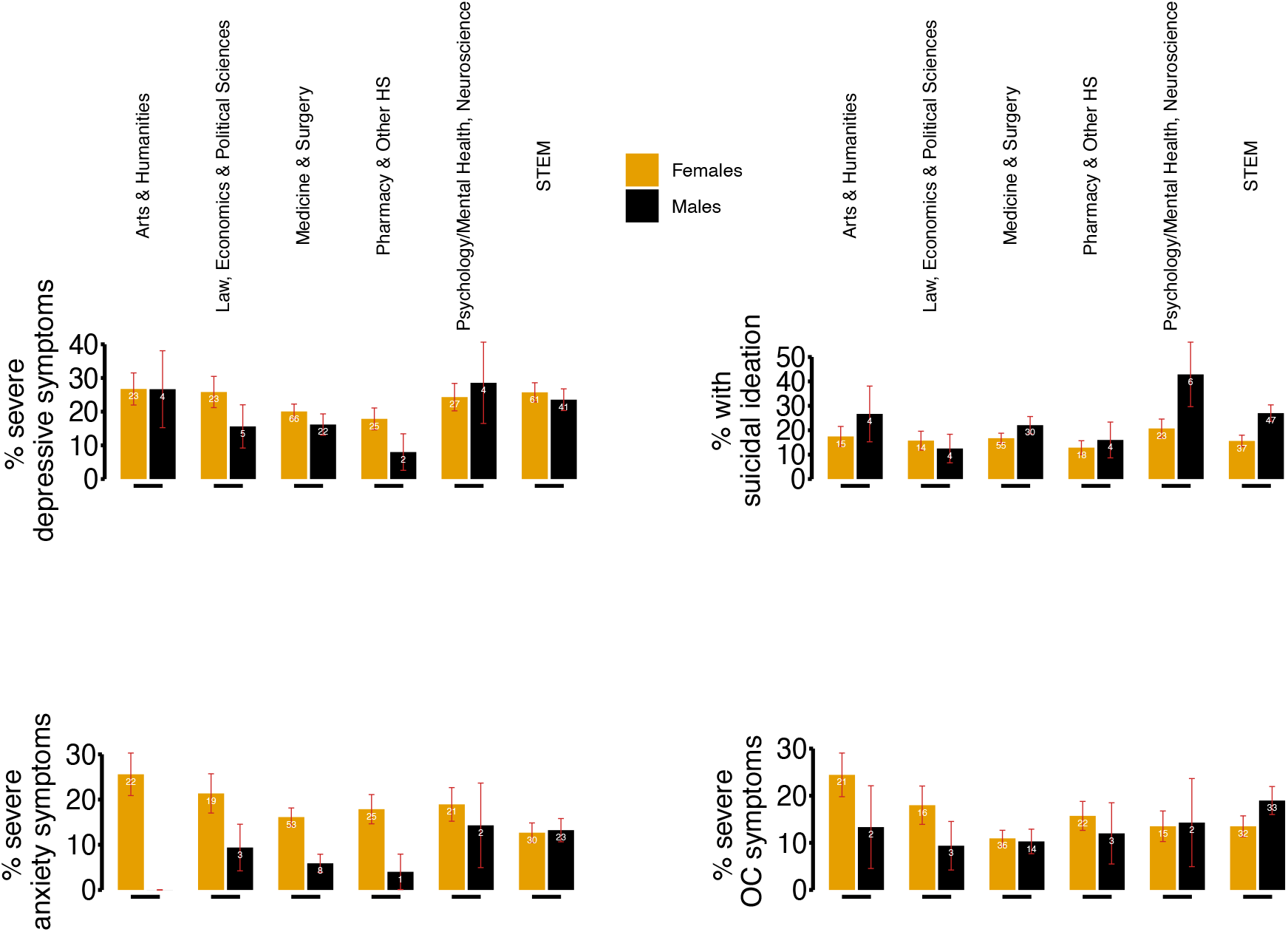
Frequency of severe depressive, anxiety symptoms and suicidal ideation among students per field of study and gender. OC = Obsessive-Compulsive (symptoms); HS = Health Sciences; STEM = Science, Technology, Engineering and Mathematics. Orange bars represent female participants, black bars male participants. In red inside each bar: error bar; the number on top of the orange/black bar represents the absolute number (white) of participants of a specific field of study experiencing severe symptoms. Absolute numbers and frequencies are also detailed in Table 1.

### Significant variables associated with severe symptoms of depression

We defined several binomial regression models to assess the contribution of demographic variables and clinically relevant characteristics to a higher severity of depression, anxiety, obsessive-compulsive symptoms, and the presence of suicidal ideation.

The model that best describes the severity of depressive symptoms, measured with the Beck Depression Inventory, is reported in Table 2. We evidenced an association between high and extreme frequency of worry for one’s economic situation, with odds of having severe symptoms of depression approximately tripled (odds ratio – OR – for high frequency of worry: 3.11 [1.88 – 5.15], p < 0.001; OR for extreme frequency of worry: 3.51 [1.53 – 7.57], p < 0.01). In addition, severe depressive symptoms are associated with having followed at least one diet in life (OR = 1.77 [1.31 – 2.39], p < 0.001). The largest association is between suicidal ideation and depression severity, with the former increasing the odds of depression severity by approximately nine times (OR = 9.29 [6.71–12.86], p < 0.001). We also evinced some factors associated with a lower likelihood of manifesting severe symptoms: either commuting or living off-site (residence outside of Padova by more than 50Km), as well as whether enrollment in the study took place before the first ever COVID-19-related lockdown (March to May 2020). Both variables approximately halved the chances of participants reporting severe symptoms of depression. The performance of the model was good, with R2 = 0.310.

### Significant variables associated with severe anxiety symptoms and obsessive-compulsive symptoms

For each of the other measures (i.e., anxiety severity, obsessive-compulsive symptoms severity, and presence of suicidal ideation), we report two models: one agnostic with respect to the BDI-II total score and its subscales score, the other instead leveraged this information to produce a better fit of the data to the model. The reason for describing these two models (i.e., with or without the information produced by the BDI-II) is that the Beck Anxiety Inventory, Obsessive-Compulsive Inventory – Revised, and the presence of suicidal ideation (measured with item number nine of the Beck Depression Inventory – II) is that their scores might be influenced by the presence of depression/depressive symptoms (i.e., the higher the burden of depressive symptoms, the higher the likelihood of these scores being elevated).

We found a variable associated with severe anxiety symptoms: suicidal ideation. Its presence increases twofold (Table 3A, OR = 2.09 [1.49 – 2.93], p < 0.001, Table 2) the odds of severe symptoms. When leveraging the score of the BDI-II and its subscales, the previous association is no longer significant and superseded by the BDI-II somatic subscale score (OR = 1.09 [1.07 – 1.12], p < 0.001 – meaning that for each point on the scale, the odds of severe anxiety symptoms are increased by approximately 9%). However, for both models, performance was very low (R^2^ = 0.034 or 0.07, respectively).

**Table 3.**
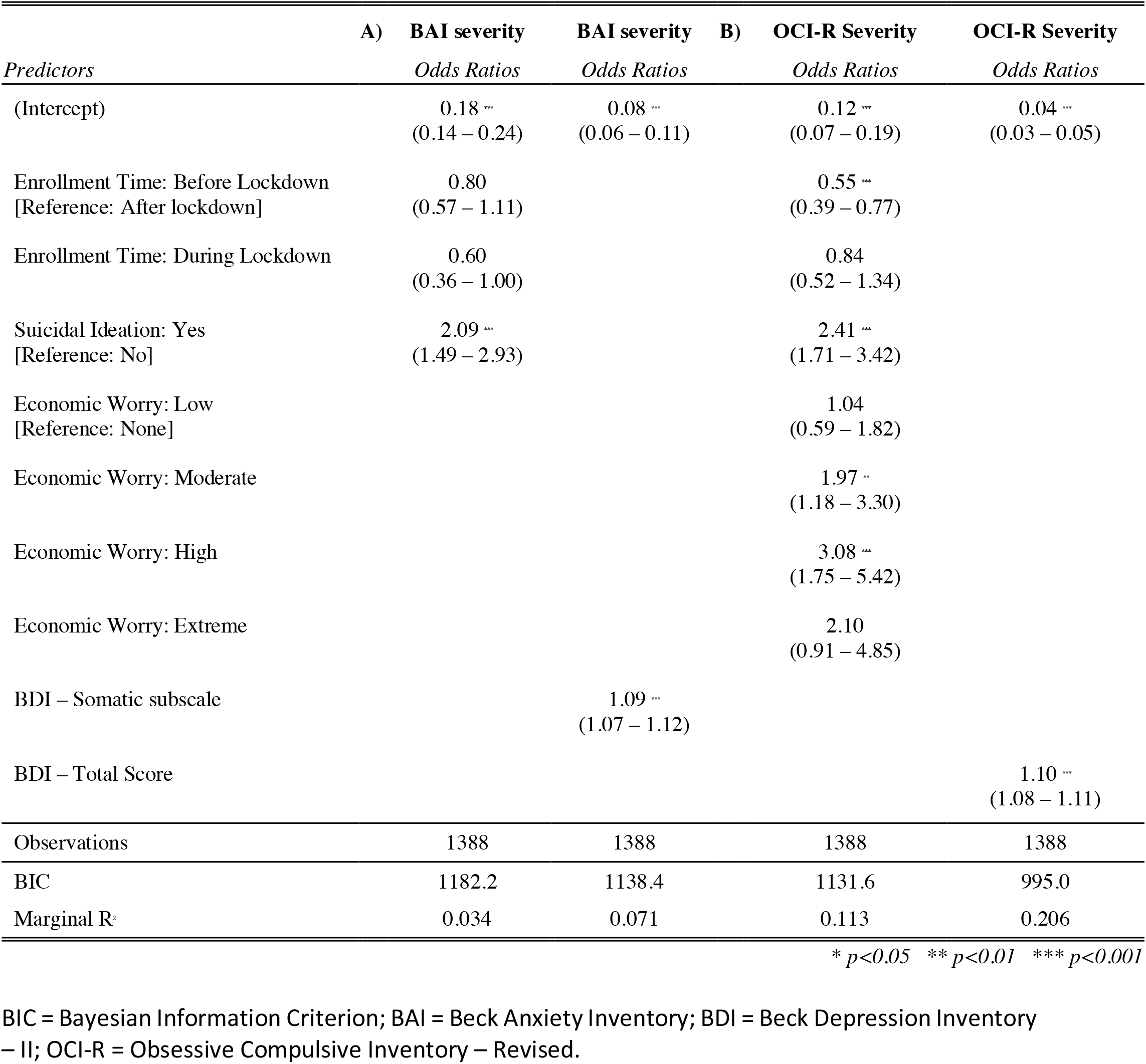
Regression models for severe anxiety or obsessive-compulsive symptoms.

Regarding obsessive-compulsive symptoms, we found that frequency of economic worry increased by two- to three-fold the odds of severe symptoms (Table 3B, moderate frequency OR = 1.97 [1.18–3.30], p < 0.01; high frequency OR = 3.08 [1.75 – 5.42], p < 0.001; extreme frequency was not significant in this model); moreover, enrollment before lockdown decreases the odds of severe symptom by 50% (OR = 0.55 [0.39 – 0.77], p < 0.001). Finally, suicidal ideation increased the severity of symptoms with an OR of 2.41 ([1.71 – 3.42], p < 0.001). The model fit was low (R^2^ = 0.113). However, considering the total score of BDI-II in the model provides a better fit (although still low, R^2^ = 0.2) for each additional point on the scale, the odds of symptoms increased by 10% ([1.08 – 1.11], p < 0.001)

### Significant variables associated with current suicidal ideation

A large number of variables were significantly associated with suicidal ideation: we show a protective role of enrollment before lockdown (Table 4, OR = 0.56 [0.4 – 0.78], p < 0.001) and marginally significant protection linked to enrollment during lockdown (OR = 0.58, [0.36 – 0.95], p < 0.05). The absence of a history of diagnosed mental disorder was found to be a significant protective factor with a large effect (OR = 0.28 [0.2 – 0.4], p < 0.001).

**Table 4.**
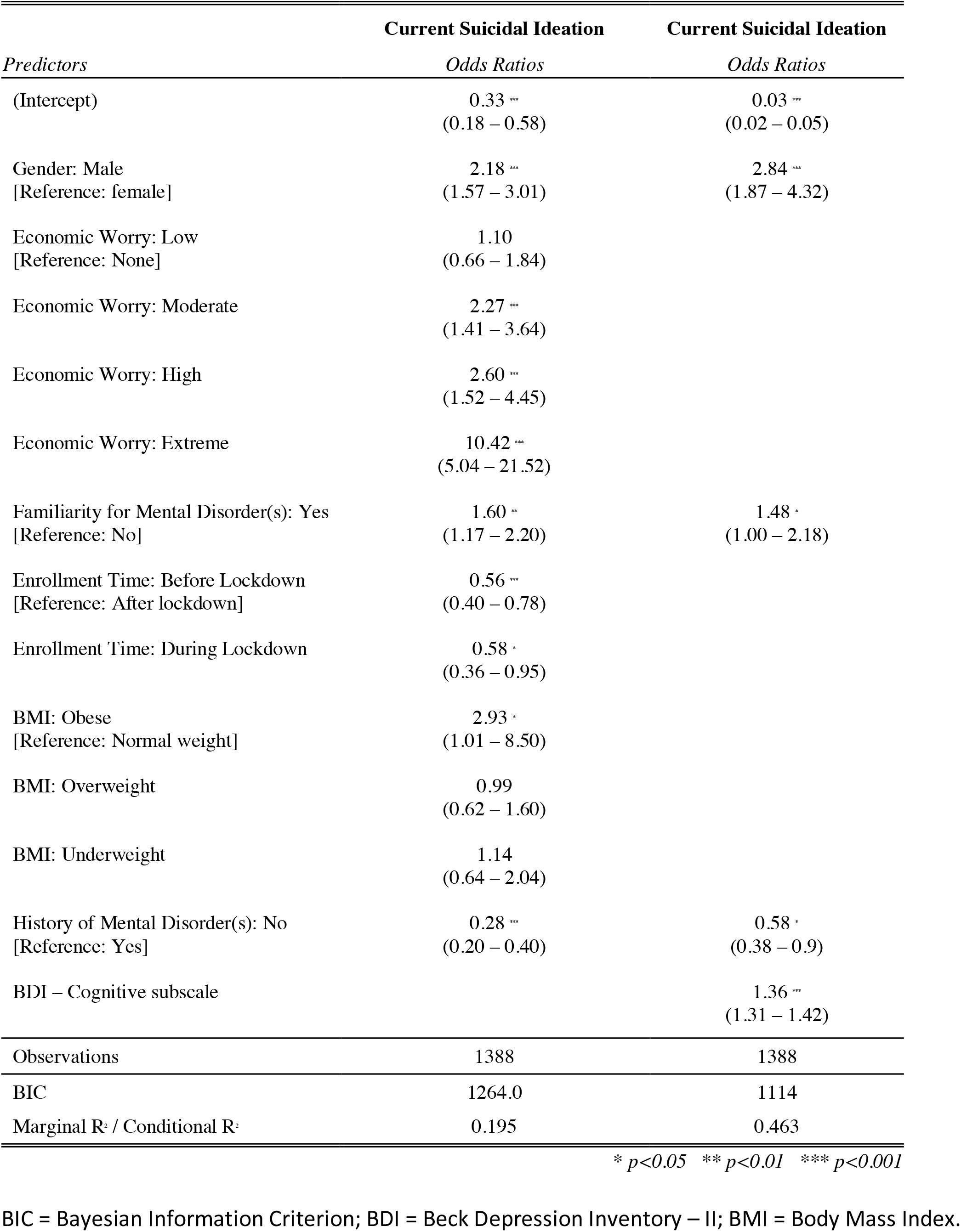
Regression models for current suicidal ideation.

Regarding risk factors, the male gender increased the odds of suicidal ideation approximately two times (OR = 2.18 [1.57 – 3.01], p < 0.001). Furthermore, the frequency of economic worry was identified as a possible risk factor, with increasing odds of suicidal ideation, as the frequency varied from moderate to extreme (for the latter, a ten-fold increase was observed [5 – 21.5], p < 0.001). A familial history of mental disorder was associated with 60% higher odds ([1.17 – 2.2], p < 0.01). A statistically marginal significance of BMI was shown (being obese, OR = 2.93 [1.01 – 8.5], p < 0.05). When covarying for BDI-II scores, we found that the score to the cognitive subscale of depression (thus measuring the variables related to cognitive distortions – in the scoring of the subscale, in this specific case, item 9 pertaining to suicidal ideation was excluded due to collinearity with the outcome measure) increased the odds of suicidal ideation by 36% for each additional point to the subscale ([1.31 – 1.42], p < 0.001). No history of mental disorders maintained a significant protective role, although the effect size was scaled down (OR = 0.58 [0.38 – 0.9], p < 0.05). However, marginal significance was reached for familiarity with mental disorder (if present, OR = 1.48 [1.00 – 2.18], p < 0.05). It is noteworthy that the size of the male gender effect increased, with the odds of suicidal ideation in males being 2.84 times higher than in females ([1.87 – 4.32], p < 0.001). The R^2^ for this model was 0.46 and should be considered highly acceptable (considering the volatility of the outcome measure).

### Comparison between follow-up and drop-out samples

Six months after enrollment, 557 participants completed a second wave of questionnaires (Table 5). We compared the samples of participants who participated in follow-up or dropped out. Follow-up completers were significantly more likely to have a familiar history of mental disorders, major in Medicine & Surgery, and be enrolled before the lockdown. No other statistically significant variables were evidenced.

**Table 5.**
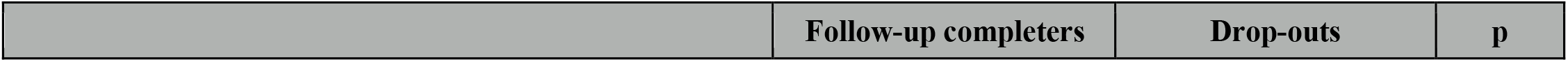
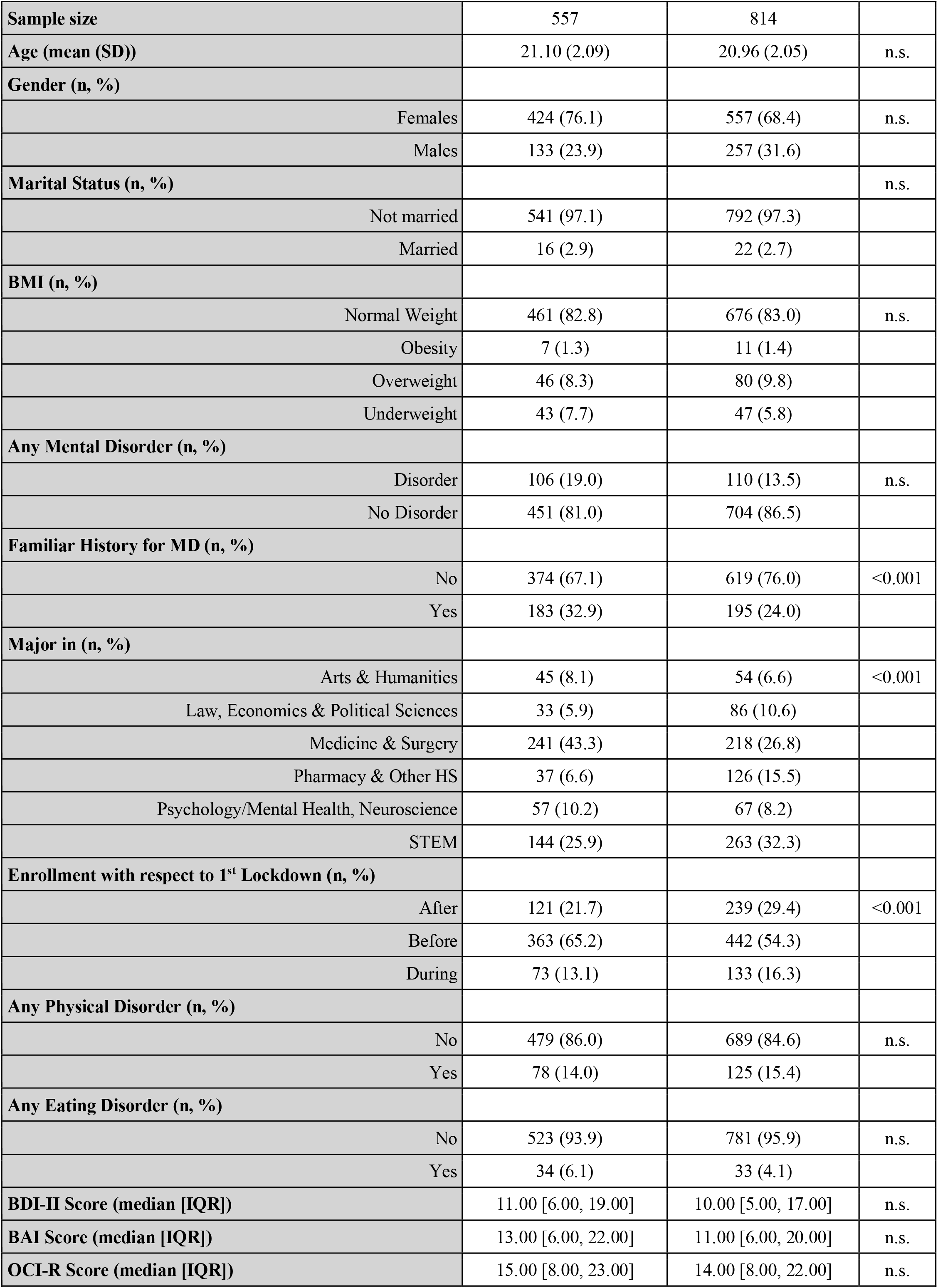
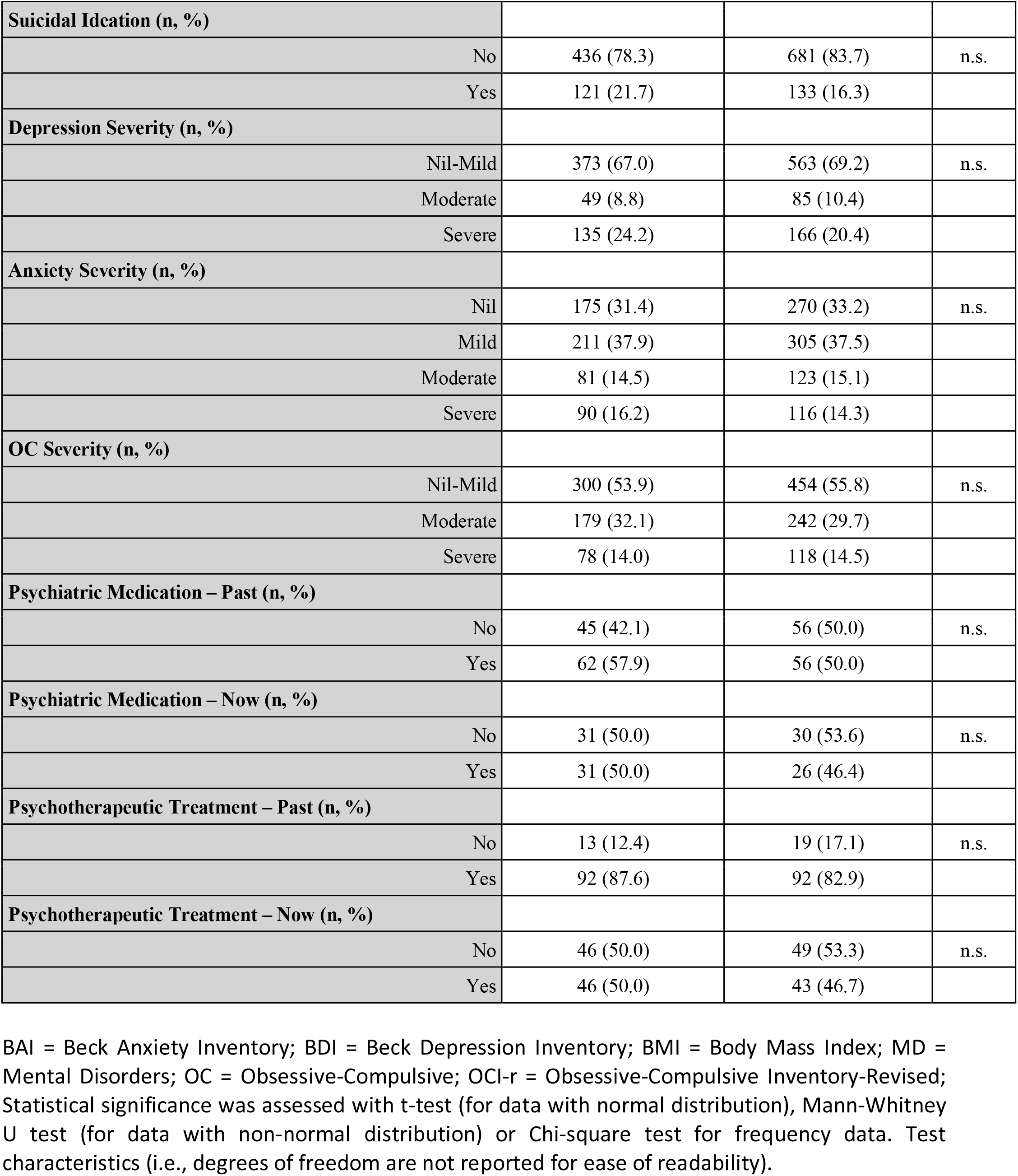
Comparison between follow-up completers and drop-outs.

|  | **Follow-up completers** | **Drop-outs** | **p** |
| --- | --- | --- | --- |
| **Sample size** | 557 | 814 |  |
| **Age (mean (SD))** | 21.10 (2.09) | 20.96 (2.05) | n.s. |
| **Gender (n, %)** |  |  |  |
| Females | 424 (76.1) | 557 (68.4) | n.s. |
| Males | 133 (23.9) | 257 (31.6) |  |
| **Marital Status (n, %)** |  |  | n.s. |
| Not married | 541 (97.1) | 792 (97.3) |  |
| Married | 16 (2.9) | 22 (2.7) |  |
| **BMI (n, %)** |  |  |  |
| Normal Weight | 461 (82.8) | 676 (83.0) | n.s. |
| Obesity | 7 (1.3) | 11 (1.4) |  |
| Overweight | 46 (8.3) | 80 (9.8) |  |
| Underweight | 43 (7.7) | 47 (5.8) |  |
| **Any Mental Disorder (n, %)** |  |  |  |
| Disorder | 106 (19.0) | 110 (13.5) | n.s. |
| No Disorder | 451 (81.0) | 704 (86.5) |  |
| **Familiar History for MD (n, %)** |  |  |  |
| No | 374 (67.1) | 619 (76.0) | <0.001 |
| Yes | 183 (32.9) | 195 (24.0) |  |
| **Major in (n, %)** |  |  |  |
| Arts & Humanities | 45 (8.1) | 54 (6.6) | <0.001 |
| Law, Economics & Political Sciences | 33 (5.9) | 86 (10.6) |  |
| Medicine & Surgery | 241 (43.3) | 218 (26.8) |  |
| Pharmacy & Other HS | 37 (6.6) | 126 (15.5) |  |
| Psychology/Mental Health, Neuroscience | 57 (10.2) | 67 (8.2) |  |
| STEM | 144 (25.9) | 263 (32.3) |  |
| **Enrollment with respect to 1<sup>st</sup> Lockdown (n, %)** |  |  |  |
| After | 121 (21.7) | 239 (29.4) | <0.001 |
| Before | 363 (65.2) | 442 (54.3) |  |
| During | 73 (13.1) | 133 (16.3) |  |
| **Any Physical Disorder (n, %)** |  |  |  |
| No | 479 (86.0) | 689 (84.6) | n.s. |
| Yes | 78 (14.0) | 125 (15.4) |  |
| **Any Eating Disorder (n, %)** |  |  |  |
| No | 523 (93.9) | 781 (95.9) | n.s. |
| Yes | 34 (6.1) | 33 (4.1) |  |
| **BDI-II Score (median [IQR])** | 11.00 [6.00, 19.00] | 10.00 [5.00, 17.00] | n.s. |
| **BAI Score (median [IQR])** | 13.00 [6.00, 22.00] | 11.00 [6.00, 20.00] | n.s. |
| **OCI-R Score (median [IQR])** | 15.00 [8.00, 23.00] | 14.00 [8.00, 22.00] | n.s. |

|  |  |  |  |  |
| --- | --- | --- | --- | --- |
| **Suicidal Ideation (n, %)** |  |  |  |  |
|  | No | 436 (78.3) | 681 (83.7) | n.s. |
|  | Yes | 121 (21.7) | 133 (16.3) |  |
| **Depression Severity (n, %)** |  |  |  |  |
|  | Nil-Mild | 373 (67.0) | 563 (69.2) | n.s. |
|  | Moderate | 49 (8.8) | 85 (10.4) |  |
|  | Severe | 135 (24.2) | 166 (20.4) |  |
| **Anxiety Severity (n, %)** |  |  |  |  |
|  | Nil | 175 (31.4) | 270 (33.2) | n.s. |
|  | Mild | 211 (37.9) | 305 (37.5) |  |
|  | Moderate | 81 (14.5) | 123 (15.1) |  |
|  | Severe | 90 (16.2) | 116 (14.3) |  |
| **OC Severity (n, %)** |  |  |  |  |
|  | Nil-Mild | 300 (53.9) | 454 (55.8) | n.s. |
|  | Moderate | 179 (32.1) | 242 (29.7) |  |
|  | Severe | 78 (14.0) | 118 (14.5) |  |
| **Psychiatric Medication – Past (n, %)** |  |  |  |  |
|  | No | 45 (42.1) | 56 (50.0) | n.s. |
|  | Yes | 62 (57.9) | 56 (50.0) |  |
| **Psychiatric Medication – Now (n, %)** |  |  |  |  |
|  | No | 31 (50.0) | 30 (53.6) | n.s. |
|  | Yes | 31 (50.0) | 26 (46.4) |  |
| **Psychotherapeutic Treatment – Past (n, %)** |  |  |  |  |
|  | No | 13 (12.4) | 19 (17.1) | n.s. |
|  | Yes | 92 (87.6) | 92 (82.9) |  |
| **Psychotherapeutic Treatment – Now (n, %)** |  |  |  |  |
|  | No | 46 (50.0) | 49 (53.3) | n.s. |
|  | Yes | 46 (50.0) | 43 (46.7) |  |

BAI = Beck Anxiety Inventory; BDI = Beck Depression Inventory; BMI = Body Mass Index; MD = Mental Disorders; OC = Obsessive-Compulsive; OCI-r = Obsessive-Compulsive Inventory-Revised; Statistical significance was assessed with t-test (for data with normal distribution), Mann-Whitney U test (for data with non-normal distribution) or Chi-square test for frequency data. Test characteristics (i.e., degrees of freedom are not reported for ease of readability).

### Prediction of severe depressive symptoms after six months with supervised machine learning

We implemented a random forest algorithm to determine which variables at baseline could be leveraged to predict a change (or stationarity) in depressive symptoms. The algorithm was trained to predict four possible outcomes: stationarity of severe symptoms, stationarity of well-being, and improvement of symptoms or worsening symptoms: 79 participants experienced a worsening of symptoms – 45 from nil-mild symptoms to moderate symptoms or from moderate to severe; 34 from nil-mild symptoms to severe symptoms; 77 participants experienced a symptom improvement; 310 remained in a state of well-being, whereas 91 participants still experienced severe symptoms. The random forest trained with 80% of the dataset (n = 461) identified cognitive and somatic subscales scores of BDI-II, frequency of economic worry, and field of study as significant variables to classify participants into the four classes (Figure 2A). Furthermore, the interaction between the two subscales (Figure 2B) showed that the predictive ability of the algorithm to classify participants is heavily influenced by extreme scores: the chances of participants still struggling with severe symptoms reach a probability of almost 1 when both subscale scores are greater than 20; similarly, the probability of not having severe symptoms (subpanel ‘Still Ok’ of Figure 2A) reaches almost 1 when both subscale scores are less than 10. When tested on the remaining 20% (n = 96) of the sample, the overall accuracy of the model was 0.77 (95% CI: 0.67 – 0.85), which was statistically significant (p < 0.001), although with lower accuracy in predicting the participants whose symptoms worsened (balanced accuracy: 0.49) than participants stationarily well (0.85). In particular, the positive predictive value of worsened symptoms after six months of enrolment was 0 (whereas the negative predictive value was 0.89). Further details on model characteristics can be found in the supplementary appendix.

**Figure 2.**
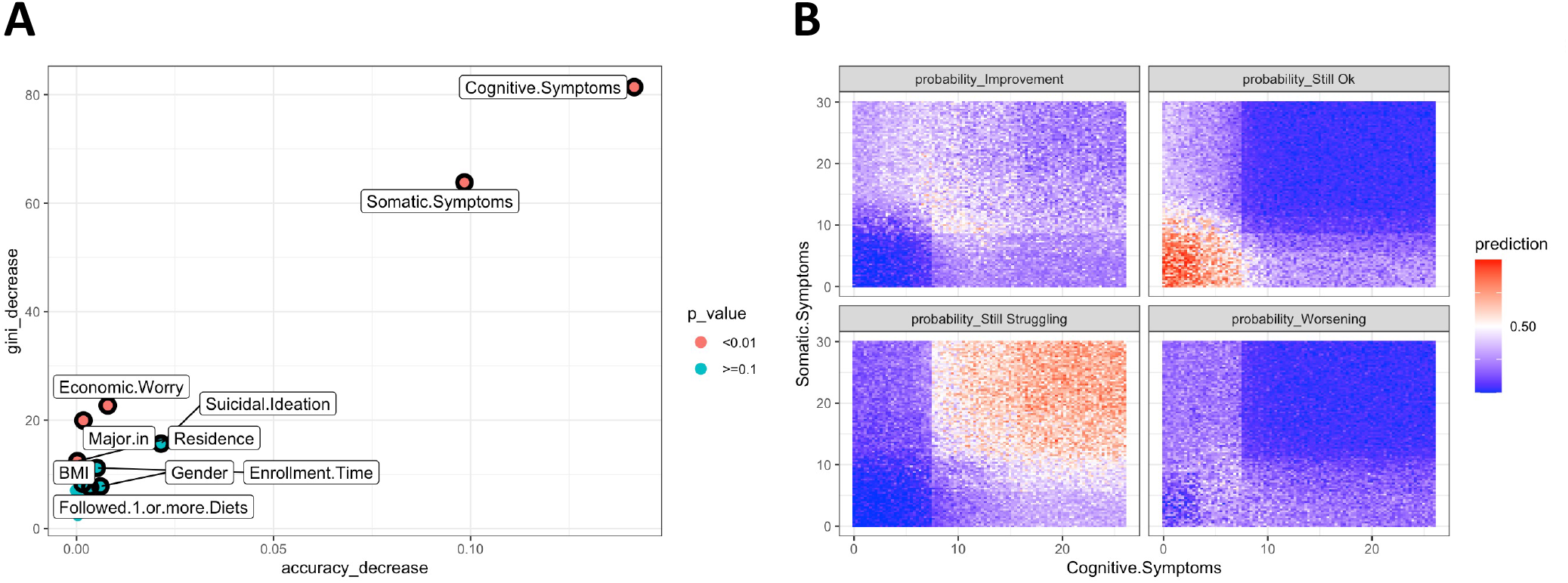
Significant variables extracted by random forest algorithms to predict severe depressive symptoms at follow-up. A) Multiway plot showing the relative importance of the top ten variables used to predict severe depressive symptoms at follow-up (six months after enrollment). Variables in red are statistically significant. A higher score in accuracy decrease or gini decrease (which is a measure of how each variable contributes to the homogeneity of the nodes and leaves) reflects the relative importance of that variable in the model (i.e., if the variable is located in the upper right corner of the plot – like cognitive symptoms – means that removing that variable from the model significantly worsen the model prediction capability). B) Predictions of the random forest of severe depressive symptoms depending on values of the BDI-II subscale scores at baseline. Participants could be classified into four classes: improvement from baseline, worsening from baseline or stability (either steadily severe symptoms or steadily well-being). For each subplot predictions ranges from 0 (deep blue) to 1 (red). For example, probability of “still struggling” reaches almost 1 (certainty) if the subscale scores at baseline are high.

### Prediction of suicidal ideation after six months with supervised machine learning

A random forest algorithm was trained to predict four possible outcomes: stationarity of suicidal ideation, stationarity of no suicidal ideation, and improvement (suicidal ideation at baseline, not at follow-up) or worsening symptoms (no suicidal ideation at baseline, present at follow-up – termed *ex novo*): 34 participants report *ex novo* suicidal ideation, whereas 48 participants reported absence of suicidal ideation at follow-up; 402 without suicidal ideation both at baseline and at follow-up; 73 reported suicidal ideation both at baseline and follow-up. The random forest trained with 80% of the data set (n = 461) identified cognitive and somatic subscale scores of BDI-II, frequency of economic worry, field of study and residence (in Padova, commuting or off-site), as significant variables to classify participants into the four classes (Figure 3A). Noteworthily, although suicidal ideation at baseline could be leveraged by the model to improve predictive capability, it was not statistically significant. Furthermore, the interaction between the two subscales of the BDI (Figure 3B) showed that the predictive ability of the classification algorithm is once again heavily influenced by extreme scores: the chances of participants not reporting suicidal ideation are high for cognitive scores below 10, with little contribution of somatic symptoms scores. The model classification performance was low for the other classes (improvement, worsening, or suicidal ideation at baseline and follow-up). When tested on the remaining 20% (n = 96) of the sample, the model overall accuracy was 0.91 (95% CI: 0.84 – 0.96), which was statistically significant (p < 0.001). However, the balanced accuracy for *ex novo* suicidal ideation was 0.5 with a negative predictive value of 0.96 but sensitivity of 0. The balance accuracy of the participants who were stationary – without suicidal ideation – was 0.92. Further details on model characteristics can be found in the supplementary appendix.

**Figure 3.**
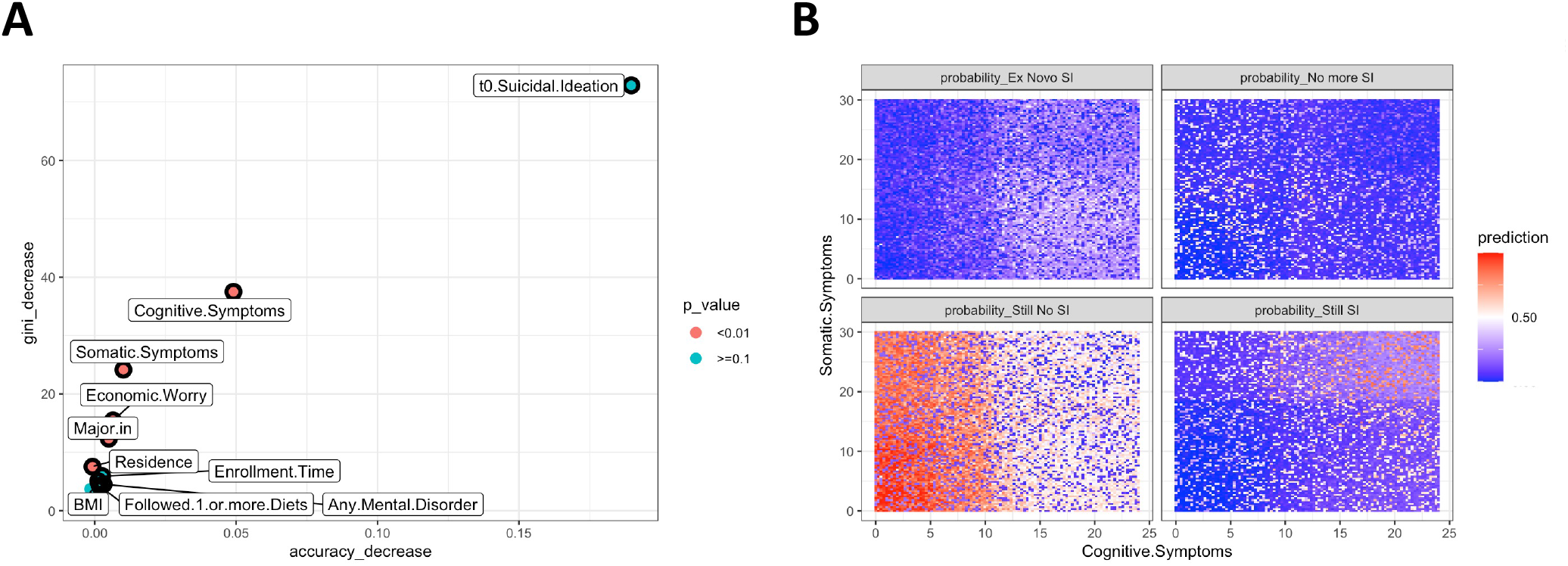
Significant variables extracted by random forest algorithms to predict suicidal ideation at follow-up. A) Multiway plot showing the relative importance of the top ten variables used to predict suicidal ideation at follow-up. Variables in red are statistically significant. A higher score in accuracy decrease or gini decrease (which is a measure of how each variable contributes to the homogeneity of the nodes and leaves) reflects the relative importance of that variable in the model. B) Predictions of the random forest of suicidal ideation depending on values of the BDI-II subscale scores at baseline.

## Discussion

Since the COVID-19 pandemic and the resulting lockdowns, mental health among students has attracted more research than ever before (Li et al., 2021; Robinson et al., 2022). Several pre-pandemic prospective studies highlighted that higher education is a risk factor for depressive symptoms, anxiety symptoms, and suicidal thoughts (Adams et al., 2021; Storrie et al., 2010).

However, there is little prospective evidence on the prediction of mental health problems. This would be the first step in a pragmatic approach to predict which students are likely to need assistance in the short, medium, and long term.

In this work, we assessed students’ depressive, anxiety, obsessive-compulsive symptoms, and suicidal ideation with their possible determinants. We collected demographic and self-report measures from 1388 Italian university students who completed questionnaires on mental health problems, collected at enrolment and after six months. We described the percentage of students suffering from severe symptoms of mental health problems; identified the factors (e.g., gender, financial situation) associated with poor mental health at baseline; and evaluated the risk factors that could predict a lack of improvement in symptoms or an increased likelihood of symptom worsening. Lastly, we show which self-report measures explained a significant part of data variance related to current and future mental health problems.

We found that the percentage of students suffering from severe depressive symptoms ranged between 22% (in females) and 17% in males. That means that in our sample 1 out of 4-6 had scored higher than the 95^th^ percentile of normative scores for the Beck Depression Inventory-II (the questionnaire used to assess depressive symptoms). Another extremely distressing finding was that 22% and approximately 20% (males and females, respectively) endorsed suicidal ideation in the two weeks before questionnaire completion. Figures for severe anxiety and obsessive-compulsive symptoms are a tad lower (severe anxiety percentages are approximately between 9% and 17%; severe OC symptoms are 14%). These findings are generally in line with previous descriptive studies on the percentages of students’ mental health problems (Robinson et al., 2022; Sheldon et al., 2021), although to some extent they are worse than expected. We consider this finding secondary to the normalization of mental health problems after the wave of COVID-19 studies, which reported that an eye-watering percentage of young adults struggled with their mental health (before, during, and after lockdowns) and on which an astounding amount of media reports were recorded. Future studies could test this hypothesis by measuring the degree of personal stigma of university students to assess whether it might have exerted a sort of Papageno effect (Etzersdorfer and Sonneck, 1998; Niederkrotenthaler et al., 2010) on more general mental health symptoms (Papageno effect, strictly speaking, is ‘the [positive] influence that the mass media can have by responsibly reporting on suicide and presenting non-suicide alternatives to crises”).

In our study, we found that people who reported high or extreme levels of worry about the economic situation were three times as likely to experience severe symptoms of depression compared to those who reported lower levels of worry. The effect between the economic situation and depressive symptoms is a bidirectional vicious cycle (Ridley et al., 2020). The results of the random forest algorithms we employed also evidenced this link, as they identified economic worry (which is, however, distinct from poverty or actual financial distress) as a useful predictor to assess future depression severity. Specifically, the algorithms showed that students with low or no concern for their economic situation had greater chances of feeling relatively well compared to their peers in financial distress. There is evidence from previous research (specifically on depressive symptoms and suicidal behaviour) that *worry* for expected financial losses is sufficient to trigger a cascade leading to depressive symptoms and suicidal thoughts (Fiksenbaum et al., 2017).

However, the association with the largest effect size (i.e., the strongest association) is between suicidal ideation and the severity of depression, the former increasing the probability of severe symptoms of depression by approximately nine times. This has to be contextualized by taking into account that suicidal ideation, as measured with the BDI, is a measure of depression itself (and a crucial diagnostic criteria for major depressive disorder, as conceptualized by the DSM-5 (Cai et al., 2021; Liu et al., 2022)).

Regarding the evaluation of the factors associated with severe symptoms of anxiety, for every one-point increase on the BDI-II somatic subscale, the odds of being severely anxious increase by approximately 9%. In relation to obsessive-compulsive symptoms, we found that every extra point on the BDI-II scale increased the chance of developing symptoms by 10%. There could be two interpretations to this finding: the fact that higher scores to the BDI-II tend to increase (as a rising tide lifts all boats) and/or both phenomena are comorbid (Jenkins et al., 2021; Norton et al., 2008; Torres et al., 2016). More studies should consider measuring obsessive, anxiety, and depressive symptoms with more than one scale to eventually unravel the contagion effects of one scale score on the other measures.

When co-varying for depression scores, some variables were significantly associated with the presence of suicidal ideation: we found that the score on the cognitive subscale of depression (which measured the variables related to cognitive distortions) increased the odds of suicidal ideation by 36% for each additional point on the subscale. Marginal significance for familiarity with mental disorders was evidenced. Whereas no history of mental disorders played a significant protective role. It is noteworthy that self-identification with the male gender increased the likelihood of suicidal ideation by 2.84 times. As reported in the literature, depressive symptoms exert a strong effect on suicidal ideation (Konick and Gutierrez, 2005; Wang et al., 2017). Regarding gender, recent evidence contrasted the view that female gender posits a greater risk of suicidal ideation (Eskin et al., 2011; Rogers and Joiner, 2017). The fact that young adult males are at higher risk of suicidal ideation has lately gained traction (Lima et al., 2021; Yarar et al., 2023). These can be better understood in light of the more extensive use of statistical tools, taking into account multiple variables simultaneously, and thus examining the corrected weights of gender on suicidal ideation (Gui et al., 2022), although cultural differences need to be thoroughly assessed, as they can shift the weights in favour of males or females (Kaggwa et al., 2023).

Lastly, we employed random forest algorithms to evince what factors at baseline were predictive of a change in depressive symptoms (i.e., still severe symptoms, worsening symptoms, improvement of symptoms, still no severe symptoms) and suicidal ideation (i.e., still suicidal ideation, *ex novo* suicidal ideation, improvement of suicidal ideation or still no suicidal ideation). For both measures, the supervised machine learning algorithms identified the scores on the cognitive and somatic subscales of BDI, the frequency of economic worry, the field of study, and the residence at baseline (the latter only for suicidal ideation) as significant variables to leverage to predict the change (or stability) of depressive symptoms. Noteworthily, suicidal ideation at baseline could be leveraged by the machine to predict suicidal ideation after six months, but it did not yield a strictly statistically significant role. To our knowledge, there have been several studies implementing machine learning models leveraging previous suicidal ideation to predict future suicidal ideation (Benjet et al., 2022; Liao et al., 2022; Malone et al., 2021), although timeframes from baseline to follow-up did not coincide with our six-month timeframe. Substantially, previous suicidal ideation is an important factor, posing a greater risk of future suicidal ideation and suicidal behavior (Bafna et al., 2022), but in our analysis did not reach canonical statistical significance (although it should be taken into account that the machine learning model would lose a great proportion of its accuracy without data from previous suicidal ideation – Figure 3A). The use of machine learning in future studies could help find new associations between the variables, although limitations must be considered: when dealing with rare outcomes (e.g., new onset of suicidal ideation after several months from baseline), algorithms learn better to predict the more common outcomes (e.g., still no suicidal ideation). We think this unbalance in the data (i.e., outcomes of stability being over-represented with respect to changes in symptomatology) can be effectively dealt with through multicentric collaborations. This approach would simultaneously address two tricky situations: cross-cultural / geographic differences and small sample sizes.

### Strengths and Limitations

The main strengths of this study are the prospective design, the analysis of various psychological dimensions through the compilation of validated questionnaires, the large cohort size, and the careful acquisition of demographic variables. However, it should be considered that self-reporting questionnaires are not diagnostic instruments, and therefore they might not detect relevant changes in the mental health of participating subjects. Another limit concerns follow-up: approximately 1/3 of the individuals agreed to participate in the study again six months after enrolment. Although the baseline differences between follow-up completers and drop-outs are negligible in terms of the outcomes of interest, the prospective evidence presented herein must be taken with caution and further supported by more research.

## Data Availability

The anonymised dataset and code for analysis can be retrieved at https://www.researchgate.net/profile/Nicola-Meda-2

## Author contributions

Conceptualisation: NM, SP, FV, CN

Methodology: NM, SP, FV, CN

Investigation: NM, SP, FV, CN

Software: PR, NM

Formal analysis: NM

Visualisation: NM

Supervision: FV, CN

Writing – original draft: NM

Writing – review & editing: NM, SP, FV, CN

## Acknowledgments

The authors would like to extend gratitude to all students who, without monetary incentives, gave their time and essential contribution to the study. We would like to extend our thanks to all the Professors and Department directors who helped us spread awareness of the study. Lastly, we would like to thank Dr Irene Slongo, Dr Giulia Fattorini, Dr. Christian Durante for help with participants recruitment. We thank Eng. Michele Gasparini (Evoé, Padova, Italy) for hosting the kick-off meeting of this project.

