## Supplementary Material for "Frequency and machine learning predictors of depressive, anxiety, obsessive-compulsive symptoms, and suicidal ideation among university students"

**Running title: Mental health problems among university students**

**Authors:** Nicola Meda^1^ MD, Susanna Pardini^2^ MSc, Paolo Rigobello^3^, Francesco Visioli^3,*^ PhD, Caterina Novara^4^ PhD

**Affiliations:**

^1^ Department of Neuroscience, University of Padova, Via Giustiniani 5, Padova, Italy

^2^ Department of General Psychology, University of Padova, Via Venezia 8, Padova, Italy

^3^ Department of Molecular Medicine, University of Padova, Viale G. Colombo 3, Padova, Italy

^4^ IMDEA-Food, CEI UAM + CSIC, Carr. de Canto Blanco 8, E, Madrid, Spain

* Corresponding author:

Prof. Francesco Visioli,

Department of Molecular Medicine, University of Padova,

Viale G. Colombo 3, Padova, Italy

**INDEX**

Summary of Random Forest model (on train dataset) for depressive symptoms…………………………...3

Summary of model performance on the test dataset………………………………….………………………………….7

Summary of Random Forest model (on train dataset) for suicidal ideation….……………….………………. 8

Summary of model performance on the test dataset.…………………………………………………………………. 12

**Summary of Random Forest model (on train dataset) for depressive symptoms**

```
Number of trees: 500
No. of variables tried at each split: 3

OOB estimate of  error rate: 29.72%
Confusion matrix:
                 Improvement Still Ok Still Struggling Worsening class.error
Improvement               30        8               28         1  0.55223881
Still Ok                   3      237                0        11  0.05577689
Still Struggling          23        1               49         1  0.33783784
Worsening                  9       51                1         8  0.88405797
```

**Distribution of minimal depth**

The plot below shows the distribution of minimal depth among the trees of your forest. Note that:

- the mean of the distribution is marked by a vertical bar with a value label on it (the scale for it is different than for the rest of the plot),
- the scale of the X axis goes from zero to the maximum number of trees in which any variable was used for splitting.

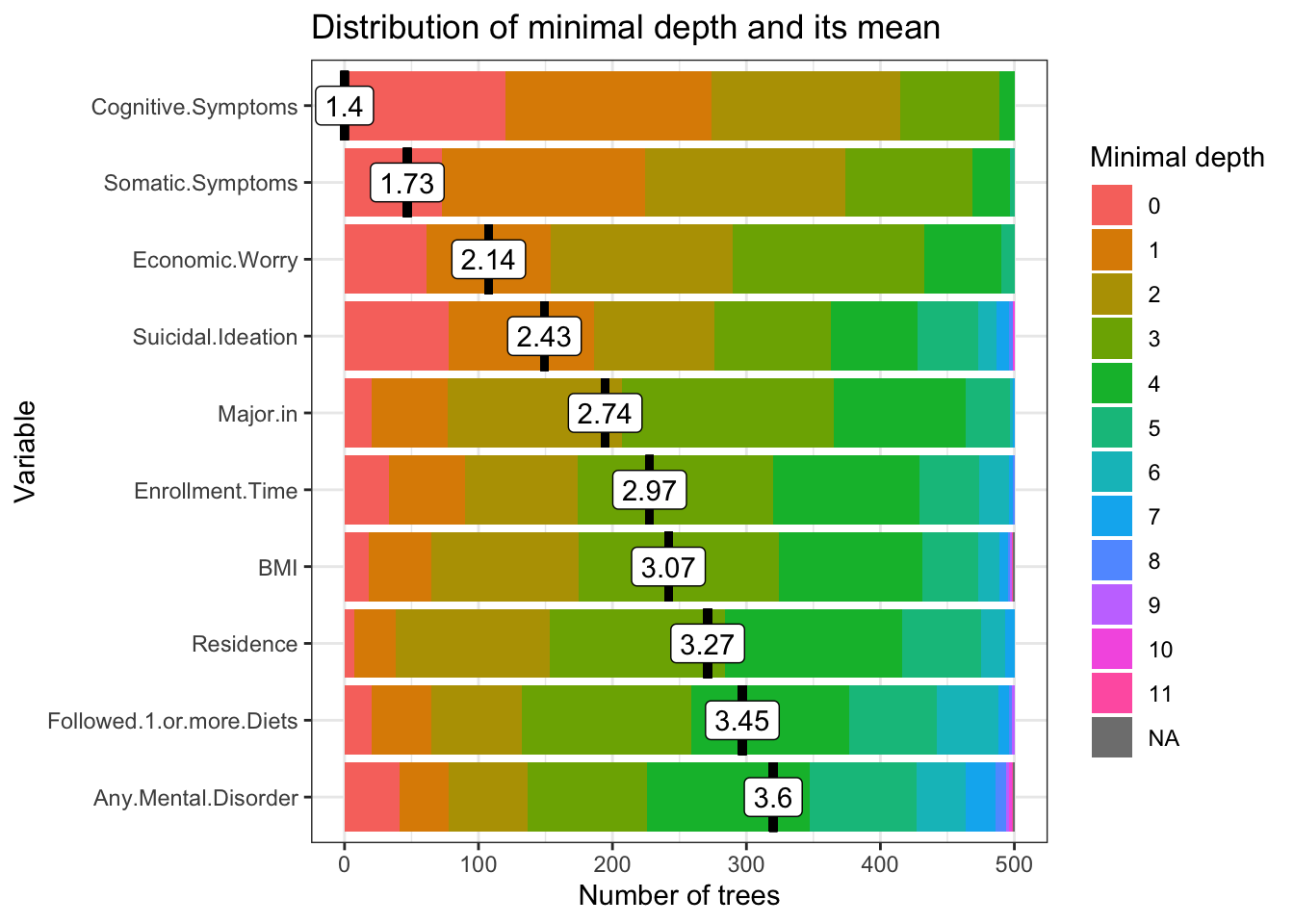

Minimal depth for a variable in a tree equals to the depth of the node which splits on that variable and is the closest to the root of the tree. If it is low than a lot of observations are divided into groups on the basis of this variable

**Multi-way importance plot**

The multi-way importance plot shows the relation between three measures of importance and labels 10 variables which scored best when it comes to these three measures (i.e. for which the sum of the ranks for those measures is the lowest).

The multi-way importance plot focuses on three importance measures that derive from the structure of trees in the forest:

- mean depth of first split on the variable,
- number of trees in which the root is split on the variable,
- the total number of nodes in the forest that split on that variable.

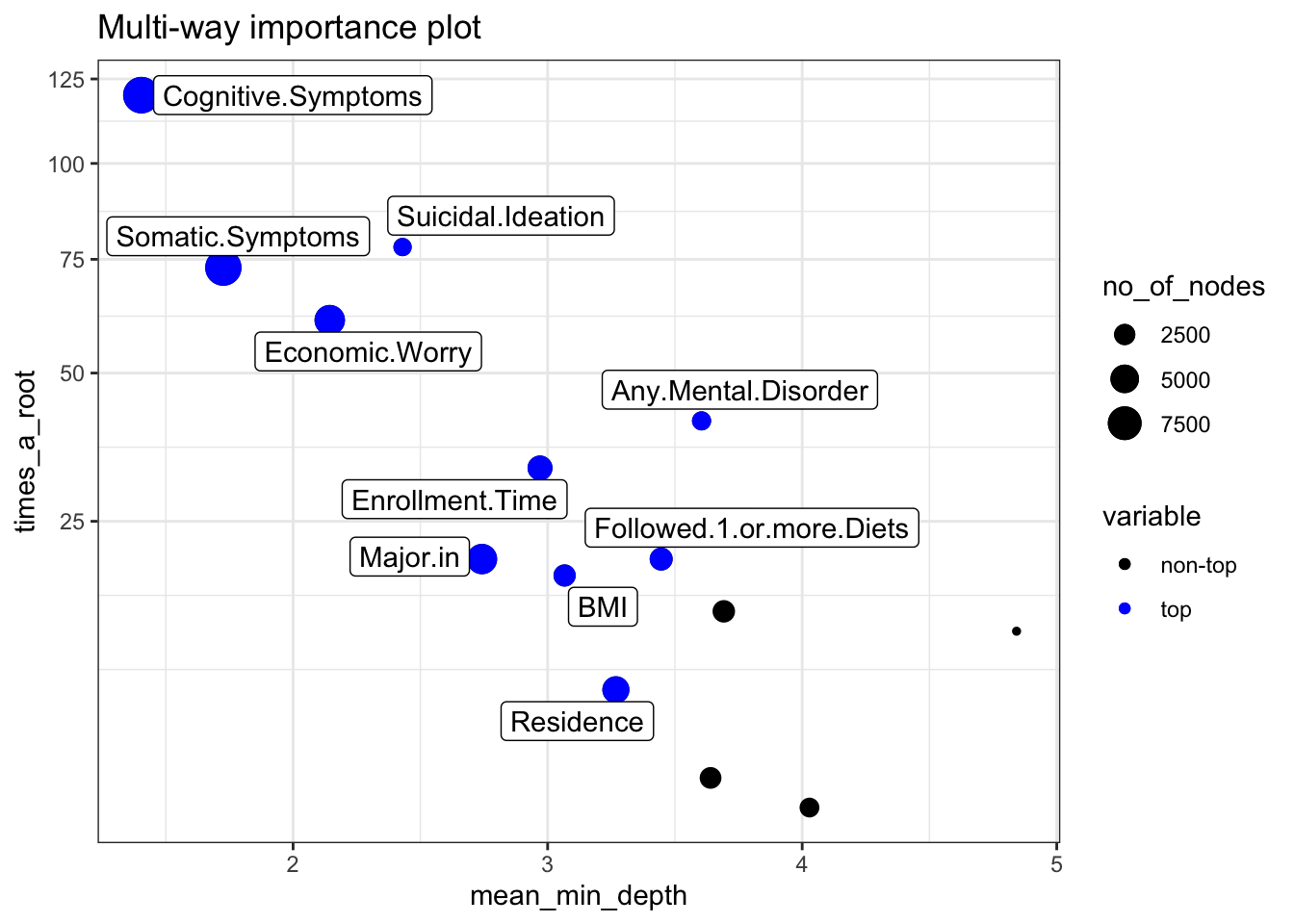

### Variable interactions

#### **Conditional minimal depth**

The plot below reports 30 top interactions according to mean of conditional minimal depth – a generalization of minimal depth that measures the depth of the second variable in a tree of which the first variable is a root (a subtree of a tree from the forest). In order to be comparable to normal minimal depth 1 is subtracted so that 0 is the minimum.

For example value of 0 for interaction x:y in a tree means that if we take the highest subtree with the root splitting on x then y is used for splitting immediately after x (minimal depth of x in this subtree is 1). The values presented are means over all trees in the forest.

Note that:

- the plot shows only 30 interactions that appeared most frequently,
- the horizontal line shows the minimal value of the depicted statistic among interactions for which it was calculated,
- the interactions considered are ones with the following variables as first (root variables): Cognitive.Symptoms, Somatic.Symptoms, Economic.Worry, Major.in, Enrollment.Time, Suicidal.Ideation, Residence, Followed.1.or.more.Diets, BMI, Gender, Any.Physical.Disorder, Familiar.History.for.MD, Any.Mental.Disorder, Any.Eating.Disorder and all possible values of the second variable.

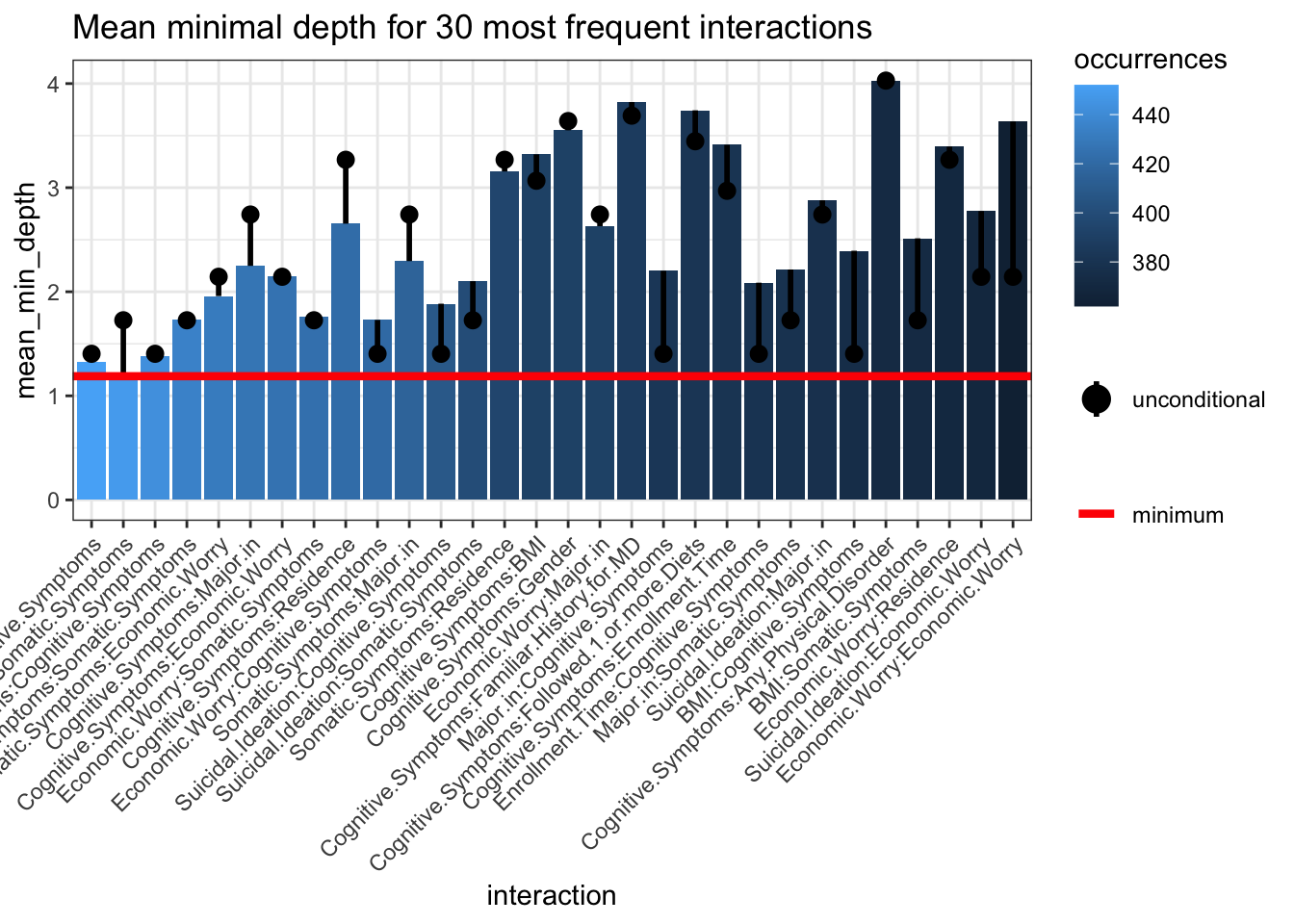

**Summary of model performance on the test dataset**

Confusion Matrix and Statistics

| Prediction | Improvement | Still Ok | Still Struggling | Worsening |
| --- | --- | --- | --- | --- |
| Improvement | 4 | 0 | 5 | 0 |
| Still Ok | 2 | 58 | 0 | 8 |
| Still Struggling | 4 | 0 | 12 | 2 |
| Worsening | 0 | 1 | 0 | 0 |

Statistics by Class:

|  | Improvement | Still Ok | Still Struggling | Worsening |
| --- | --- | --- | --- | --- |
| Sensitivity | 0.40 | 0.98 | 0.70 | 0 |
| Specificity | 0.94 | 0.72 | 0.92 | 0.98 |
| Pos Pred Value | 0.44 | 0.85 | 0.66 | 0 |
| Neg Pred Value | 0.93 | 0.96 | 0.93 | 0.89 |
| Prevalence | 0.10 | 0.61 | 0.17 | 0.10 |
| Detection Rate | 0.04 | 0.60 | 0.12 | 0 |
| Detection Prevalence | 0.09 | 0.70 | 0.18 | 0.01 |
| Balanced accuracy | 0.67 | 0.85 | 0.81 | 0.49 |

**Summary of Random Forest model (on train dataset) for suicidal ideation**

```
Number of trees: 500
No. of variables tried at each split: 3

OOB estimate of  error rate: 16.49%
Confusion matrix:
            Ex Novo SI No more SI Still No SI Still SI class.error
Ex Novo SI           1          0          30        0 0.967741935
No more SI           0         20           0       23 0.534883721
Still No SI          1          0         326        0 0.003058104
Still SI             0         22           0       38 0.366666667
```

**Distribution of minimal depth**

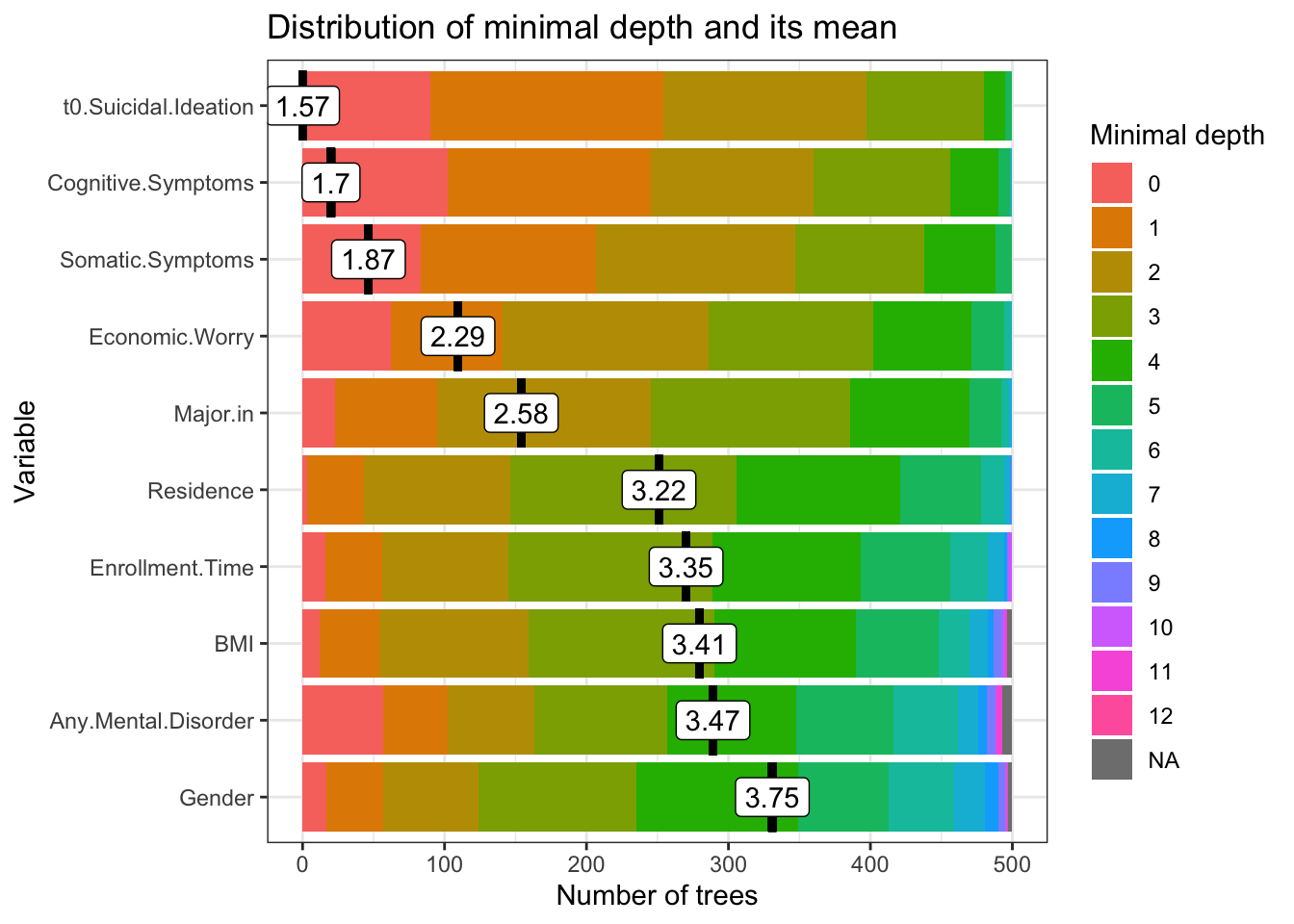

Minimal depth for a variable in a tree equals to the depth of the node which splits on that variable and is the closest to the root of the tree. If it is low than a lot of observations are divided into groups on the basis of this variable

**Multi-way importance plot**

The multi-way importance plot shows the relation between three measures of importance and labels 10 variables which scored best when it comes to these three measures (i.e. for which the sum of the ranks for those measures is the lowest).

The multi-way importance plot focuses on three importance measures that derive from the structure of trees in the forest:

- mean depth of first split on the variable,
- number of trees in which the root is split on the variable,
- the total number of nodes in the forest that split on that variable.

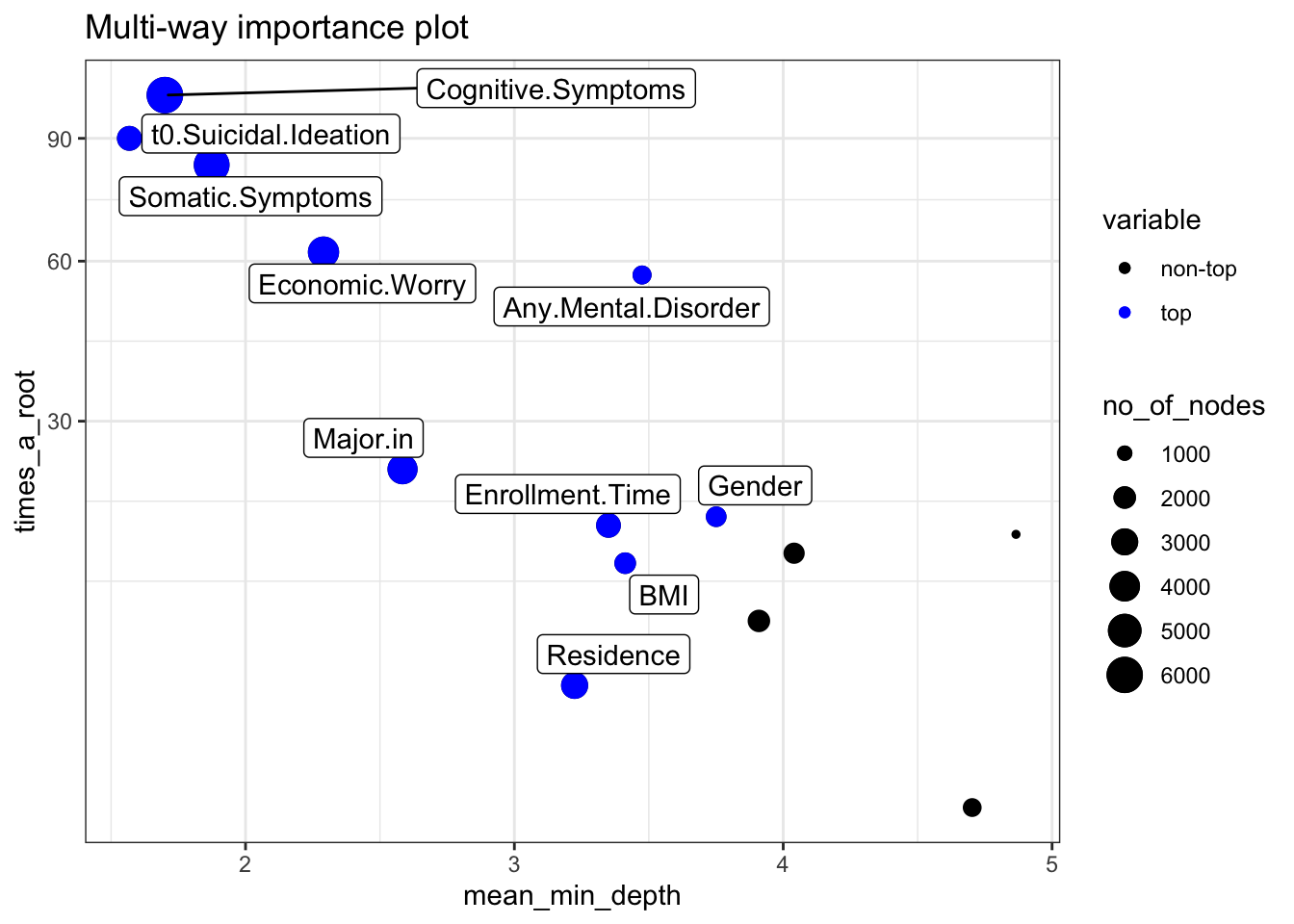

**Conditional minimal depth**

The plot below reports 30 top interactions according to mean of conditional minimal depth – a generalization of minimal depth that measures the depth of the second variable in a tree of which the first variable is a root (a subtree of a tree from the forest). In order to be comparable to normal minimal depth 1 is subtracted so that 0 is the minimum.

For example value of 0 for interaction x:y in a tree means that if we take the highest subtree with the root splitting on x then y is used for splitting immediately after x (minimal depth of x in this subtree is 1). The values presented are means over all trees in the forest.

Note that:

- the plot shows only 30 interactions that appeared most frequently,
- the horizontal line shows the minimal value of the depicted statistic among interactions for which it was calculated,
- the interactions considered are ones with the following variables as first (root variables): Cognitive.Symptoms, t0.Suicidal.Ideation, Somatic.Symptoms, Economic.Worry, Major.in, Enrollment.Time, Residence, BMI, Any.Mental.Disorder, Followed.1.or.more.Diets, Gender, Familiar.History.for.MD, Any.Physical.Disorder, Any.Eating.Disorder and all possible values of the second variable.

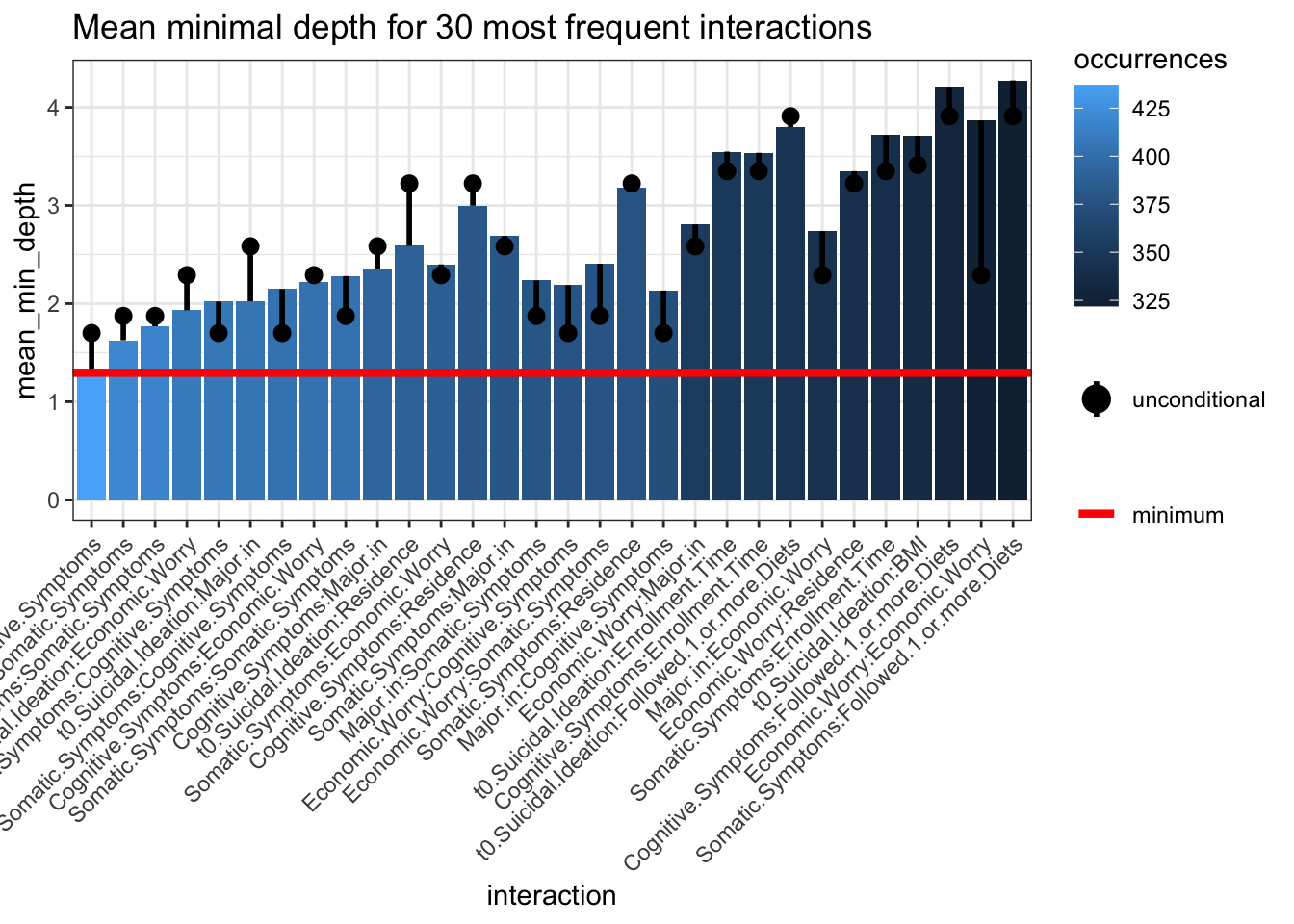

**Summary of model performance on the test dataset**

Confusion Matrix and Statistics

| Prediction | Ex novo SI | No More SI | Still No SI | Still SI |
| --- | --- | --- | --- | --- |
| Ex novo SI | 0 | 0 | 0 | 0 |
| No More SI | 0 | 2 | 0 | 2 |
| Still No SI | 3 | 0 | 75 | 0 |
| Still SI | 0 | 3 | 0 | 11 |

Statistics by Class:

|  | Ex novo SI | No More SI | Still No SI | Still SI |
| --- | --- | --- | --- | --- |
| Sensitivity | 0 | 0.40 | 1 | 0.84 |
| Specificity | 1 | 0.97 | 0.85 | 0.96 |
| Pos Pred Value | NaN | 0.5 | 0.96 | 0.78 |
| Neg Pred Value | 0.96 | 0.96 | 1 | 0.97 |
| Prevalence | 0.03 | 0.05 | 0.78 | 0.13 |
| Detection Rate | 0 | 0.02 | 0.78 | 0.11 |
| Detection Prevalence | 0 | 0.04 | 0.81 | 0.14 |
| Balanced accuracy | 0.5 | 0.68 | 0.92 | 0.90 |
